# ECG-Derived Synthetic Left Ventricular Mechanics Predict Mortality After Acute Myocardial Infarction and Fatal Myocardial Infarction in the General Population

**DOI:** 10.64898/2026.10.21.26366159

**Authors:** Douglas Corsi, John Furst, Ankush D. Jamthikar, Sajeev Singh, Gabriela M M Paixão, Antonio Luiz P. Ribeiro, Jagat Narula, Bernard R. Chaitman, Jeffrey Carson, Naveena Yanamala, Partho P. Sengupta

**Author notes:** Corresponding Author: Partho P. Sengupta, MD, DM, Chair, Department of Cardiology, Mayo Clinic Health System, 1400 Bellinger Street, Eau Claire, WI 54703. Joint first authors and equal contribution (Douglas Corsi, John Furst).

## Abstract

**Background:** Left ventricular (LV) mechanics predict outcomes after acute myocardial infarction (AMI). Recovering them from the 12-lead electrocardiogram (ECG) could enable risk assessment wherever an ECG is acquired.

**Objectives:** To test whether ECG-derived synthetic LV mechanics predict mortality after AMI and myocardial infarction (MI)-specific mortality in the general population beyond the GRACE score and ejection fraction (EF).

**Methods:** Synthetic peak systolic (s′) and early diastolic (e′) mitral annular velocities and mitral annular plane systolic excursion (MAPSE) were derived from the ECG by a validated generative adversarial network, applied without retraining and blinded to outcome. We studied an AMI cohort (n=1,065; Cox regression) and a population cohort (n=219,564; Fine-Gray models), testing incremental value over GRACE, three artificial intelligence (AI)-ECG reduced-EF models, and measured EF.

**Results:** In the AMI cohort (109 deaths; median follow-up 1.44 years), all three parameters predicted all-cause mortality, but only synthetic e′ remained independent of clinical covariates (HR=0.90 per 1 cm/s; 95% CI 0.84-0.98) or GRACE (HR=0.90; 0.83-0.97). Among 1,017 patients discharged alive (61 deaths), e′ improved discrimination beyond GRACE (C-index 0.67 to 0.70) and remained independent of AI-ECG-predicted and measured EF (HR 0.84-0.86 per 1 cm/s; all P≤0.004). In the population cohort (817 MI deaths; median follow-up 3.42 years), all three predicted MI-specific mortality after adjustment for age, sex, and ECG abnormalities (each P<0.001), most strongly MAPSE (sHR=0.49 per 1 cm; 95% CI 0.37-0.65).

**Conclusions:** ECG-derived synthetic LV mechanics predict mortality after AMI and fatal MI in the general population, adding information beyond GRACE and EF.

## Introduction

Acute myocardial infarction (AMI) affects approximately 7 million individuals worldwide each year, with an estimated 550,000 first episodes occurring in the United States annually [1,2]. At the core of AMI pathophysiology is a highly dynamic process of myocardial dysfunction, evolving over seconds to hours as ischemia progresses from early reversible contractile abnormalities to stunning, recovery, or irreversible necrosis and scar formation, producing a clinical spectrum ranging from limited ischemic impairment to severe pump failure and cardiogenic shock [3].

Echocardiography is routinely used to assess left ventricular (LV) function during AMI and provides insight into the evolving ischemic wavefront progressing from the subendocardium toward the epicardium [4]. Because longitudinal myocardial fibers are predominantly subendocardial, abnormalities in LV longitudinal function become increasingly pronounced with greater transmural injury [5,6], and echocardiography-derived measures of LV mechanics provide prognostic information beyond conventional risk stratification in AMI [7–9]. However, echocardiographic assessment is episodic, capturing isolated snapshots within a continuously evolving ischemic process, and is often unavailable when critical decisions are made.

The 12-lead ECG, universally obtained in AMI, encodes information about myocardial mechanical function through electromechanical coupling [10,11]. We have previously utilized generative artificial intelligence to synthesize patient-specific mechanical waveforms directly from surface ECG signals with high fidelity, generating clinically relevant and directly interpretable representations of myocardial motion that can be integrated into established risk-stratification frameworks [12]. More recently, in patients undergoing transient balloon occlusion of the left anterior descending artery, we showed that ECG-derived synthetic tissue velocities decline early during acute ischemia, typically before conventional ST-segment elevation, and recover after reperfusion [13]. This provides a mechanistic basis for the surface ECG tracking ischemic LV dysfunction in real time.

Accordingly, we evaluated whether ECG-derived synthetic LV mechanical parameters provide prognostic information across two complementary settings: all-cause mortality in hospitalized AMI patients and MI-specific mortality in a large population-based cohort, and whether these measures add prognostic value beyond conventional clinical risk stratification and left ventricular ejection fraction [14,15].

## Methods

### Study Populations and Design

We previously described a generative adversarial network (GAN) for synthetic TDI, validated by correlation and agreement with echocardiography-derived TDI [12,16]. This study provides independent, multi-cohort external validation and extension of that model across two distinct cohorts (**Figure 1**): (i) a retrospective clinical cohort of hospitalized AMI patients compared with an outpatient non-AMI reference cohort, and (ii) a primary-care outpatient population cohort [17].

**Figure 1.**
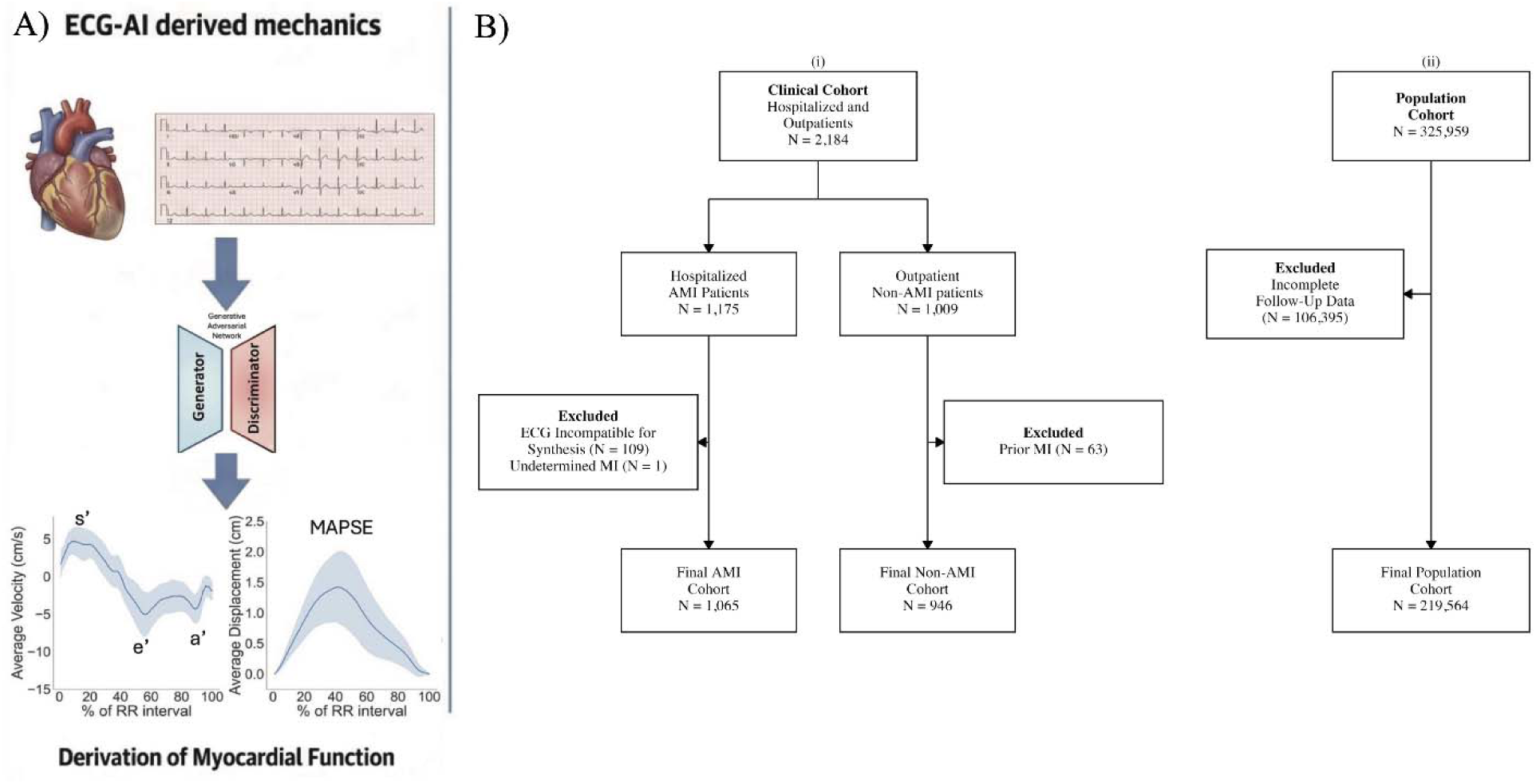
Panel A. Overview of the GAN-based ECG-to-TDI synthesis pipeline. A standard 12-lead ECG is processed through a generative adversarial network (GAN) to produce synthetic tissue Doppler imaging (TDI) mitral annular velocity waveforms (s’, e’, a’) and synthetic mitral annular plane systolic excursion (MAPSE). Panel B. Study flow diagram. Participant selection across the two study cohorts: (i) Clinical Cohort of hospitalized AMI patients and outpatient non-AMI controls, and (ii) Population Cohort from the CODE-15 Brazilian registry. Abbreviations: AMI = acute myocardial infarction; ECG = electrocardiogram; MI = myocardial infarction.

The clinical cohort comprised 2,011 patients (≥18 years) from Rutgers Robert Wood Johnson University Hospital and Newark Beth Israel Medical Center. Cases were identified from June 8, 2020, to December 28, 2024. Two prespecified groups were included: hospitalized AMI patients and outpatient non-AMI controls. Data collection was conducted as part of an NHLBI-funded research program (R01HL173998) through systematic retrospective review of electronic health records and medical charts. Deidentified data abstracted included demographics, medical history, clinical presentation, laboratory values, imaging findings, and outcomes.

AMI was classified as STEMI or NSTEMI per the Fourth Universal Definition of Myocardial Infarction [18], requiring a rise and/or fall of cardiac troponin with at least one value exceeding the 99th percentile upper reference limit accompanied by at least one of: ischemic symptoms, new ischemic ECG changes, pathological Q waves, imaging evidence of new myocardial loss or regional wall motion abnormality, or angiographic coronary thrombus. Patients were excluded if they were discharged to institutionalized care, had type 2, 4, or 5 myocardial infarction, or had coexisting terminal illness. Additionally, patients with alternative diagnoses for elevated troponin (myocarditis, pericarditis, non-ischemic cardiomyopathy, or moderate-to-severe valvular disease) and pregnant patients were excluded.

AMI status was adjudicated through two-phase blinded review: two cardiology trainees independently classified each case as STEMI, NSTEMI, or undetermined, with discordant cases resolved by an expert consultant. Of 1,175 AMI patients screened, 110 were excluded (109 for incompatible ECG sampling frequency <500 Hz; 1 undetermined), yielding 1,065 AMI patients.

Of 1,009 non-AMI patients screened, 63 were excluded for prior MI, yielding 946 controls drawn from a younger, lower-risk outpatient population to serve as a physiological reference. The non-AMI cohort was used exclusively for waveform characterization and risk-factor association analyses; all survival analyses were performed in the AMI cohort alone.

To examine the association of synthetic LV mechanics with cardiac mortality in a broad, unselected outpatient population, we analyzed a subset of the CODE-15 Brazilian registry [17]. This dataset comprises primary-care and outpatient 12-lead ECG recordings collected through the Telehealth Network of Minas Gerais, Brazil, between 2010 and 2016 (median follow-up 3.4 years; IQR 2.1–5.1). Of 325,959 participants, 106,395 were excluded for incomplete follow-up, yielding a final population cohort of 219,564 patients with complete synthetic TDI predictions and linked mortality data. Cause of death was adjudicated from national death registries and classified as cardiac (subdivided into myocardial infarction, heart failure, and other cardiac causes) or non-cardiac.

The clinical cohort was approved by the IRB of Rutgers Robert Wood Johnson Medical School (Pro2024001567 and Pro2024001026) and the population cohort by the Research Ethics Committee of the Universidade Federal de Minas Gerais (49368496317.7.0000.5149); informed consent was waived for the retrospective analysis of deidentified data. Both studies were conducted in accordance with the Declaration of Helsinki.

### ECG-Derived Synthetic LV Mechanics

Synthetic TDI parameters were generated from standard 12-lead ECGs using the previously developed and validated GAN architecture [12], applied to all cohorts without retraining and blinded to outcome. Raw tracings were segmented into individual cardiac cycles, resized to 512 timesteps, and synchronized with paired TDI recordings acquired within one hour. The GAN decoded biomechanical information embedded in the surface ECG to reconstruct myocardial velocity waveforms across the cardiac cycle at 1% temporal intervals, averaged across cycles to yield a single representative waveform per patient. Four parameters were derived: peak systolic mitral annular velocity (synthetic s′), peak early diastolic velocity (synthetic e′), peak late diastolic velocity (synthetic a′), and mitral annular plane systolic excursion (synthetic MAPSE). Lateral and septal annular values were averaged per patient. All headline analyses use the pre-procedure ECG; post-procedure tracings were used only for the paired pre-versus post-procedure comparison shown in Supplemental Figure 1.

### Analytical Aims and Outcomes

In the clinical cohort, analyses were directed at three aims. First, as a construct validity assessment, the association between synthetic and measured early diastolic velocity (e’) and established cardiovascular risk factors, including age, sex, overweight (BMI >25 kg/m²), hypertension, diabetes mellitus, dyslipidemia, smoking, obesity, chronic kidney disease, heart failure, valvular heart disease, reduced left ventricular ejection fraction (LVEF <50%), and elevated tricuspid regurgitation velocity (TRV >2.8 m/s), was examined and compared against corresponding associations with measured TDI, to evaluate whether synthetic TDI recapitulate known physiological relationships. Second, the prespecified outcome was all-cause mortality during follow-up following the index AMI hospitalization, ascertained from institutional records. Third, we assessed whether the synthetic parameters carried prognostic information incremental to the GRACE score and left ventricular systolic function, in patients discharged alive after the index admission.

In the population cohort, the prespecified outcome was cause-specific MI mortality, with mortality attributable to heart failure, other cardiac causes, and non-cardiac causes treated as competing events, enabling assessment of disease-specific mortality risk.

### Statistical Analysis

Continuous variables are reported as median (inter-quartile range, IQR) and categorical variables as counts (percentage). We used the Mann-Whitney U test to compare continuous variables between groups and the chi-square test or Fisher exact test to compare categorical variables between groups. Average waveform morphology using synthetic waveforms (TDI and displacement) for non-MI controls, AMI survivors, and AMI non-survivors was plotted across the cardiac cycle (expressed as percentage of RR interval), with shaded areas representing ±1 standard deviation around each group’s mean waveform.

Pooled MI and non-MI cohorts were analyzed using univariate and multivariable logistic regression to assess whether synthetic e’ independently differentiated infarcted from non-infarcted individuals after adjustment for clinical and echocardiographic factors. Parallel models using measured e’ were fit for comparison. Odds ratios with 95% confidence intervals are reported.

In the clinical AMI cohort, Kaplan-Meier curves stratified by tertiles were constructed for each synthetic parameter and compared by log-rank test. Univariable Cox proportional hazards models were fit for each parameter individually. To assess robustness to potential confounding by clinical risk factors, each synthetic parameter was additionally modeled separately in multivariable Cox models retaining candidate covariates (age, chronic kidney disease, atrial fibrillation, coronary territory, and prior cardiac arrest) significantly associated with all-cause mortality on univariable analysis. Model discrimination was quantified by the Harrell C-index, and the proportional-hazards assumption verified using Schoenfeld residuals. Collinearity among the synthetic parameters was assessed using generalized variance inflation factors, which were below 2.5 for every term in all multivariable models.

To evaluate incremental prognostic value beyond the GRACE 2.0 risk score, nested Cox models were compared using the likelihood-ratio test in the full AMI cohort and, separately, in the post-discharge cohort (patients discharged alive after the index admission) over the full available follow-up. Hazard ratios for the synthetic parameters are reported per 1 cm/s, or per 1 cm for MAPSE, so that a hazard ratio below 1 denotes higher mortality at lower values; synthetic e′ was additionally modeled as tertiles (2 df) with the highest tertile as reference, using cut points derived in the full AMI cohort and applied unchanged to the post-discharge cohort. The GRACE 2.0 score was available in 1,058 of the 1,065 AMI patients and in 1,010 of the 1,017 patients discharged alive.

In the population cohort, MI-specific mortality was analyzed by Fine-Gray subdistribution hazard regression with non-MI mortality as competing events; cumulative incidence functions were estimated using the Aalen-Johansen estimator and compared across tertiles using Gray’s test. A parallel sensitivity analysis used all-cause cardiac mortality as the outcome with non-cardiac mortality as the competing event. Each synthetic parameter was additionally modeled separately with adjustment for available demographic and ECG variables, including age, sex, and conventional ECG abnormalities (right and left bundle branch block, ST–T abnormality, atrial fibrillation, and first-degree atrioventricular block).

Ejection fraction was represented in four ways in the AMI cohort: three open-source AI-ECG models, applied without retraining and each dichotomized at a predicted probability of 0.5: AiTiA [19] and HeartWise [20], which predict a concurrent LVEF <50%, and the AI-ECG HFrEF risk model of Pan et al. [21], which predicts incident heart failure with reduced ejection fraction (LVEF ≤40%) over a 1–5 year horizon [21], together with measured echocardiographic LVEF <50% as the reference standard. In the population cohort only the AI-ECG HFrEF risk model of Pan et al. could be applied, because AiTiA cannot be run at population scale and HeartWise was trained using CODE-15 data. CODE-15 contributed to the development of neither AiTiA nor the model of Pan et al., so the population-cohort comparison is external for both [21]. Incremental discrimination conferred by the synthetic TDI parameters was assessed by likelihood-ratio test in the AMI cohort, with the χ² contributed by each index expressed as a proportion of the full model χ², and by paired Wolbers competing-risk concordance in the population cohort, which treats competing deaths as non-events rather than censoring them and so stays within the Fine-Gray framework used for the primary analysis; concordance was evaluated over the full CODE-15 follow-up and confidence intervals for ΔC come from paired concordance influence functions, so every comparison is made within the same participants. All analyses were hypothesis-driven. Analyses used R 4.5.2 and Python 3.8.18; two-sided P<0.05 was significant.

## Results

### Baseline Characteristics of the Clinical Cohort

The AMI cohort (n=1,065; 54% STEMI, 46% NSTEMI) had a median age of 65 years (IQR: 57–75), was predominantly male (68%), and carried a high cardiovascular risk burden, with hypertension (72.7%), prior PCI (68.5%), and prior MI (42.1%) most prevalent; 66.1% had a high GRACE 2.0 score (>140), and overall mortality was 10.2% (**Table 1**). Compared with non-AMI controls, AMI patients demonstrated higher rates of ST-T abnormalities (39.1% vs. 17.8%) and atrial fibrillation (7.1% vs. 5.8%), alongside lower synthetic s′ (6.1 vs. 6.7 cm/s) and MAPSE (1.6 vs. 1.8 cm), reflecting greater myocardial dysfunction (Supplemental Table 1).

**Table 1.**
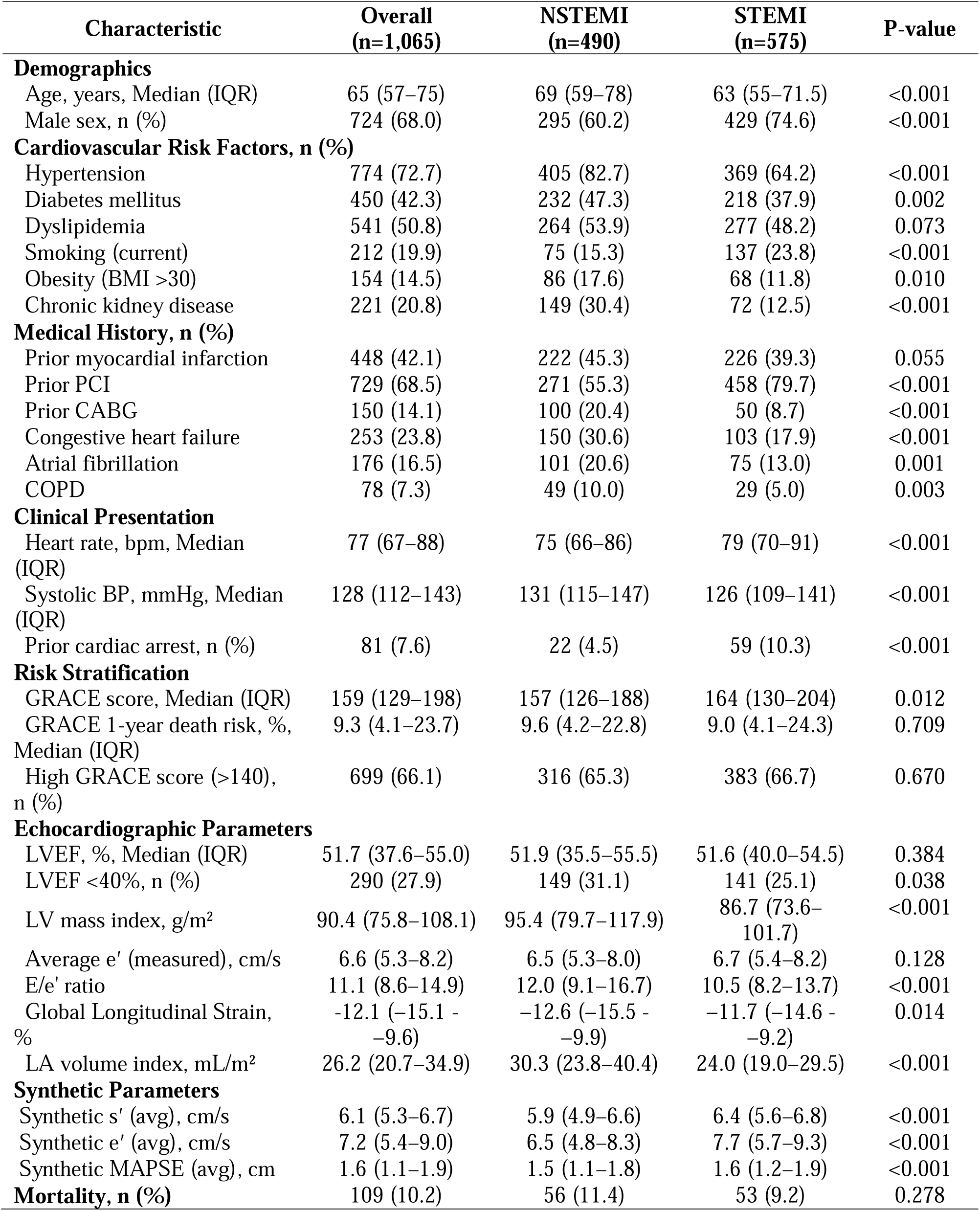
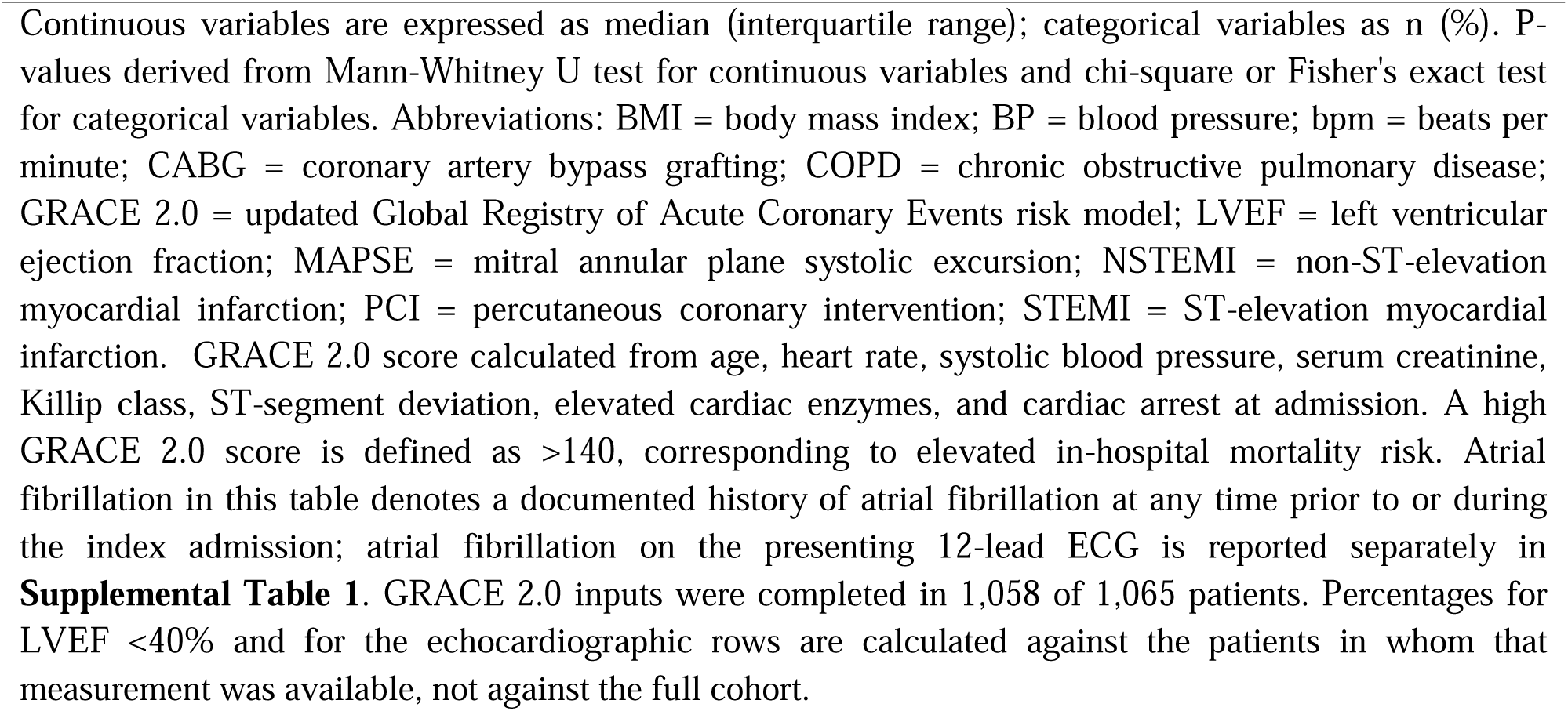
Baseline Characteristics of the Clinical AMI Cohort.

Group-averaged synthetic velocity and displacement waveforms for non-AMI and AMI patients are shown in **Figure 2**. Compared with non-MI controls (**Figures 2a** and **2b**), AMI survivors exhibited attenuated amplitudes (Figures 2c and 2d), while AMI non-survivors demonstrated further attenuation of both systolic and diastolic amplitudes (**Figures 2e** and **2f**), with a stepwise reduction in peak systolic displacement across the three groups.

**Figure 2.**
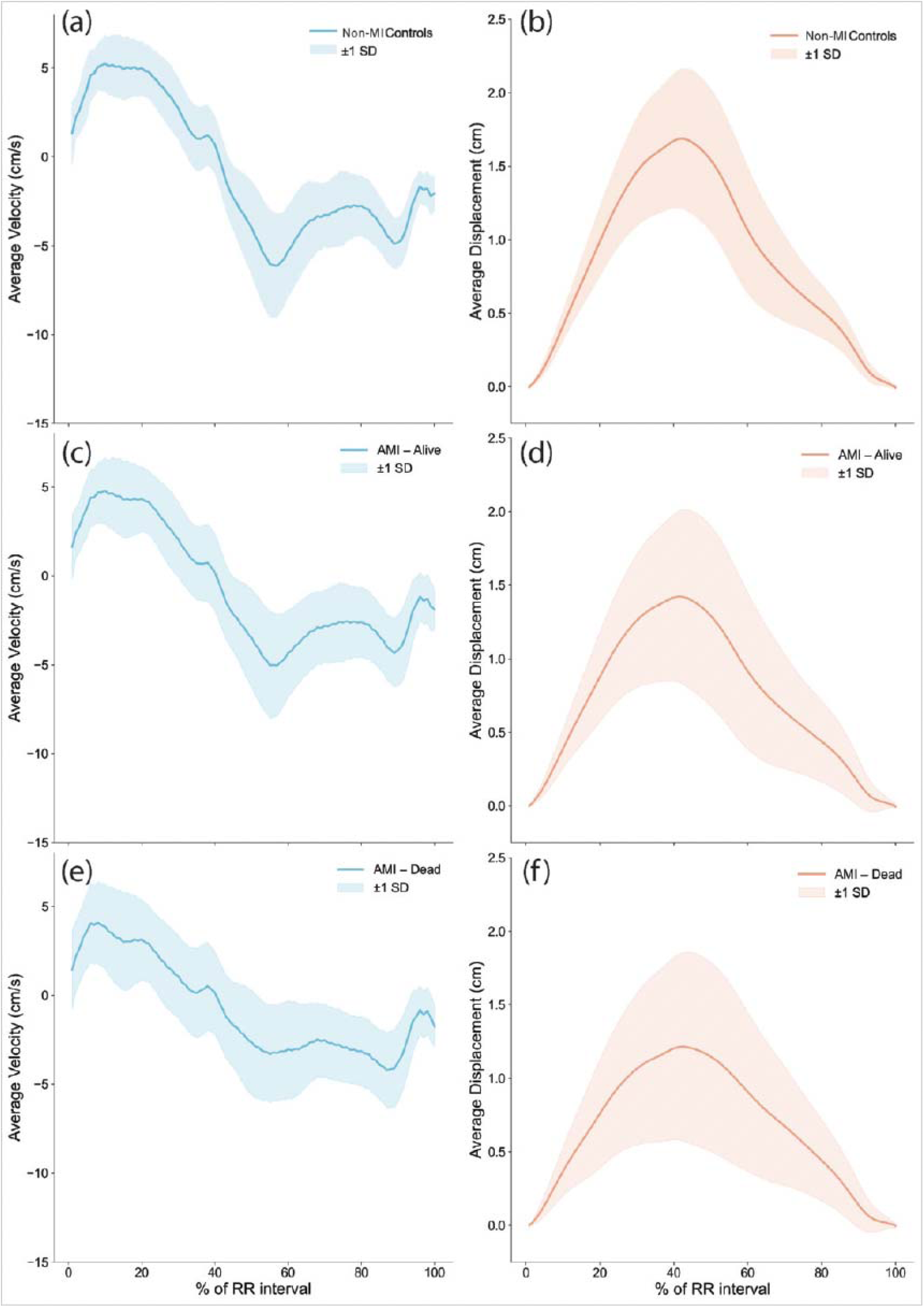
Synthetic mitral annular velocity and displacement profiles across the cardiac cycle in non-MI controls, AMI survivors, and AMI non-survivors. Group-averaged synthetic TDI velocity (left panels, cm/s) and displacement (right panels, cm) waveforms across the cardiac cycle (% RR interval) for non-MI controls (a, b), AMI survivors (c, d), and AMI non-survivors (e, f). Shaded bands represent ±1 SD. A stepwise attenuation of systolic and early diastolic amplitudes is observed from controls through survivors to non-survivors.

### Measured vs. Synthetic TDI Associations with Risk Factors and AMI

Both measured and synthetic e′ demonstrated consistent and directionally concordant univariate associations with all 13 cardiovascular risk factors (Supplemental Figure 1; ORs 0.71 to 0.98). On multivariable logistic regression, AMI independently predicted reduced both measured and synthetic e′ after adjustment for cardiovascular risk factors (Supplemental Table 2).

### Prognostic Value of Synthetic TDI in the AMI Cohort

During a median follow-up of 1.44 years (IQR 0.48-2.60), 109 patients (10.2%) died. Kaplan-Meier analysis demonstrated a significant survival gradient across tertiles of all three synthetic parameters (Figure 3, panels a–c, log-rank P<0.01 for all, tertile boundaries in **Supplemental Table 3**). On univariable Cox regression (**Supplemental Table 4**), each parameter independently predicted all-cause mortality: synthetic s′ (HR=0.80 per 1 cm/s; 95% CI 0.71–0.92; P=0.001), synthetic e′ (HR=0.85 per 1 cm/s; 95% CI 0.79–0.92; P<0.001), and synthetic MAPSE (HR=0.57 per 1 cm; 95% CI 0.41–0.80; P=0.001).

**Figure 3.**
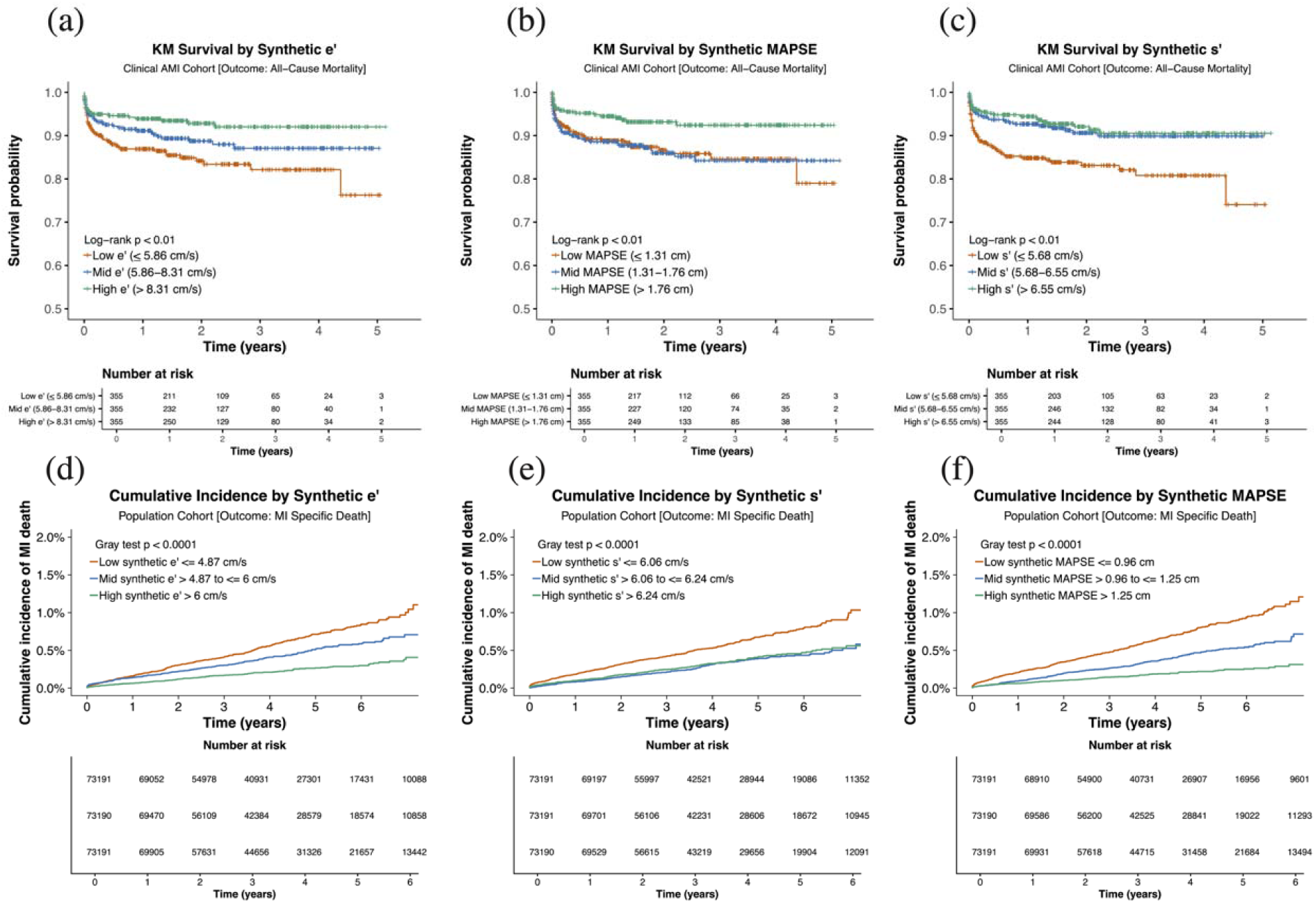
All-cause and myocardial infarction–specific mortality stratified by tertiles of synthetic tissue Doppler imaging indices. Kaplan-Meier survival curves for all-cause mortality in the Clinical Cohort (left panels) and Aalen-Johansen cumulative incidence functions for cause-specific MI mortality with non-MI mortality as competing events in the Population Cohort (right panels), stratified by tertiles of synthetic s′, synthetic e′, and synthetic MAPSE (tertile boundaries in **Supplemental Table 3**). Between-group comparisons used the log-rank test (all P<0.01) and Gray’s test (all P<0.0001). Tick marks denote censored observations. Abbreviations: e′ = early diastolic mitral annular velocity; MAPSE = mitral annular plane systolic excursion; MI = myocardial infarction; s′ = peak systolic mitral annular velocity; TDI = tissue Doppler imaging.

In the multivariable Cox models incorporating candidate covariates that were significantly associated with all-cause mortality on univariable analysis (P<0.05: age, chronic kidney disease, atrial fibrillation, prior cardiac arrest, and coronary territory) (**Supplemental Table 4**), synthetic e′ remained the only independently significant predictor (HR=0.90; 95% CI 0.84–0.98; P=0.012), whereas synthetic s′ (HR=0.90; 95% CI 0.78–1.04; P=0.143) and MAPSE (HR=0.77; 95% CI 0.54–1.09; P=0.144) were not significant.

### Prognostic Association Benchmarked Alongside GRACE

In the AMI cohort (N=1,058 with an available GRACE 2.0 score; 109 deaths), GRACE alone provided good discrimination for all-cause mortality (C-index=0.78; 95% CI 0.74–0.82, **Supplemental Table 5**); only synthetic e′ retained independent significance when added to GRACE and significantly improved model fit (HR=0.90; 95% CI 0.83–0.97; C-index=0.79; likelihood-ratio P=0.0058), whereas s′ (likelihood-ratio P=0.125) and MAPSE (likelihood-ratio P=0.16) did not. Because a substantial share of AMI deaths in the full cohort occurred during the index hospitalization, prior to any post-discharge risk-stratification decision point, we pre-specified the post-discharge cohort as the setting of clinical interest (N=1,017, of whom 1,010 had a GRACE 2.0 score; 61 deaths over a median of 536 days, IQR 236 to 978, Supplemental Table 5). In this cohort GRACE alone discriminated more modestly (C-index 0.67; 95% CI 0.60 to 0.74), and all three synthetic indices added independent prognostic information beyond it. Synthetic e′ again contributed most (adjusted hazard ratio 0.84 per 1 cm/s; 95% CI 0.76 to 0.93; P=0.001; likelihood-ratio P<0.001), raising discrimination from 0.67 to 0.70. The association was monotonic across categories of e′ tertiles: patients in the lowest e′ tertile (≤5.89 cm/s) had a 2.72-fold higher post-discharge mortality than those in the highest tertile (>8.33 cm/s) after adjustment for GRACE (95% CI 1.32 to 5.60; P=0.006; 2-df likelihood-ratio P=0.014; **Supplemental Table 6**, Supplemental Methods).

### Prognostic Association Benchmarked Alongside AI-ECG Models Predicting Reduced Ejection Fraction

In the same post-discharge cohort (N=1,017; 61 deaths over a median follow-up of 536 days), we investigated whether synthetic e′ adds to AI-ECG predictions of reduced ejection fraction from AiTiA and HeartWise (concurrent LVEF <50%) and from the model of Pan et al. (incident LVEF ≤40% over a 1–5 year horizon), each dichotomized at a predicted probability of 0.5, and measured echocardiographic LVEF <50% as the reference standard (available in 997 patients). Synthetic e′ was independently associated with death in every model: adjusted hazard ratio per 1 cm/s 0.86 (95% CI 0.77 to 0.95; P=0.004) beyond an AiTiA-predicted LVEF <50%, 0.84 (0.76 to 0.93; P=0.001) beyond a Pan et al.-predicted LVEF ≤40%, and 0.85 (0.76 to 0.94; P=0.002) beyond a measured LVEF <50%, contributing 28 to 46% of the total likelihood-ratio χ² of each model (**Supplemental Table 12**). Synthetic s′ and MAPSE were also independently associated with death (hazard ratios 0.81 to 0.83 per 1 cm/s for s′ and 0.58 to 0.62 per 1 cm for MAPSE; all P<0.05) but added less in every comparison. The absolute-risk gradient was confined to patients in whom the ECG predicted a preserved ejection fraction (**Figure 4**).

**Figure 4.**
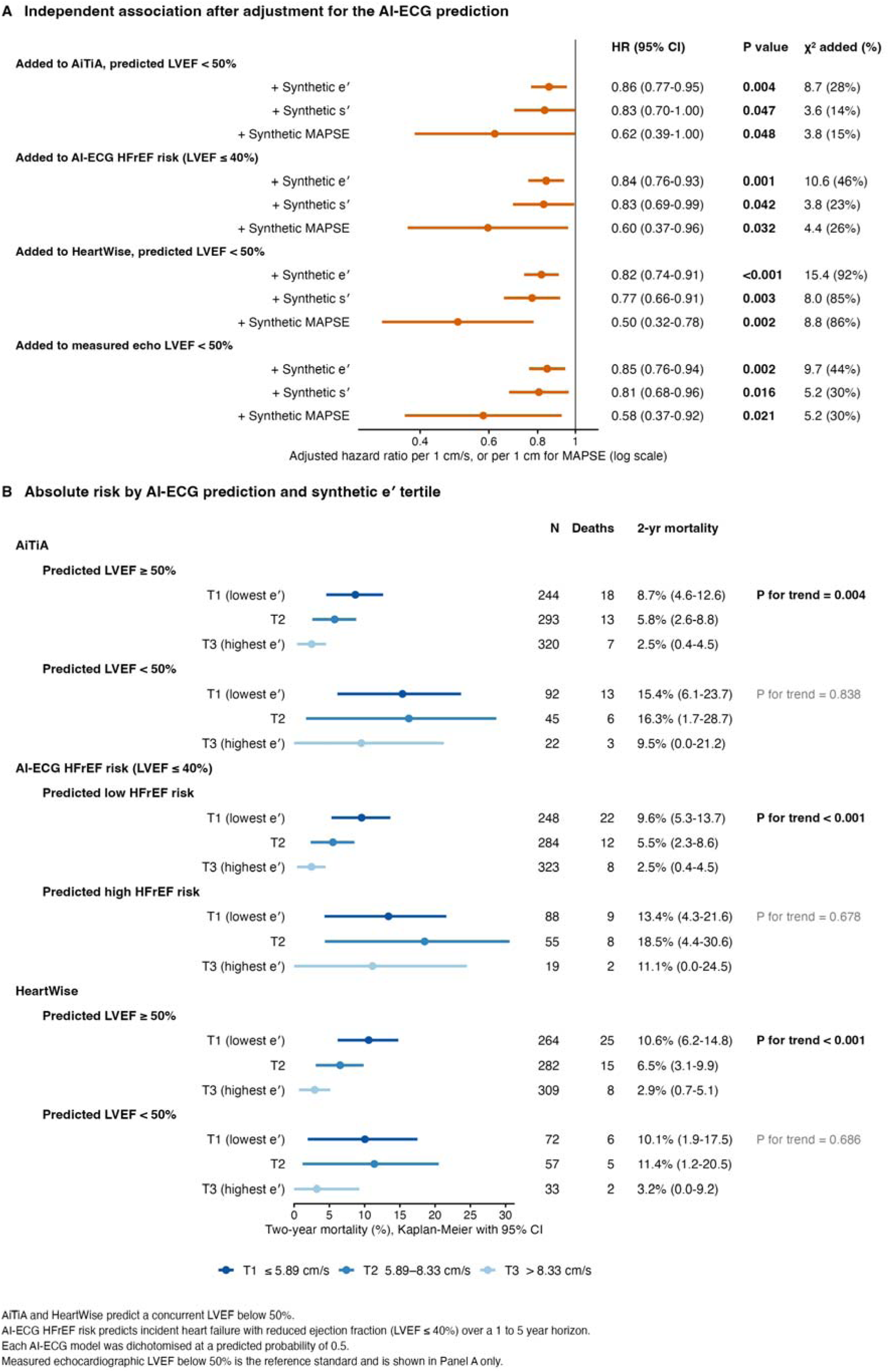
Independent association of synthetic tissue Doppler indices with mortality, and absolute risk when combined with an AI-ECG prediction of reduced ejection fraction. Among 1,017 patients who survived the index hospitalization (61 deaths over a median of 536 days), Panel A shows the adjusted hazard ratio per 1 cm/s, or per 1 cm for MAPSE, for each index when added to the ejection-fraction prediction named above it, with the likelihood-ratio χ² contributed by the index and that χ² as a percentage of the full model’s χ². AiTiA and HeartWise predict a concurrent LVEF below 50% and the model of Pan et al. predicts incident LVEF ≤40% over a 1–5-year horizon; each was dichotomized at a predicted probability of 0.5. Measured echocardiographic LVEF below 50% is shown last as the reference standard. Panel B shows two-year Kaplan–Meier mortality with 95% confidence intervals by predicted ejection fraction and tertile of synthetic e′; P values are for linear trend across tertiles within each group. Measured echocardiographic LVEF was available in 997 patients.

### Clinical Characteristics of the Population Cohort

The population cohort comprised 219,564 patients (median age 50 years, IQR 33–67; 39.6% male). During a median follow-up of 3.42 years, 10,735 deaths (4.9%) occurred: 817 (7.6%) MI-related, 887 (8.3%) heart failure, 862 (8.0%) other cardiac death, and 8,169 (76.1%) non-cardiac. Baseline ECG and synthetic TDI characteristics are presented in Supplemental Table 1; baseline characteristics stratified by vital status and adjudicated cause of death are provided in **Supplemental Table 7**.

### Synthetic LV Mechanics and MI-specific Mortality in the Population Cohort

Cumulative incidence function (CIF) analyses demonstrated graded separation of MI-mortality curves across all three parameters, with the lowest tertile carrying the highest cumulative MI-mortality incidence (**Figure 3**, panels d–f); Gray’s test was significant for all three (all P<0.0001).

On univariable Fine-Gray regression (**Supplemental Table 8**), each parameter was independently associated with MI mortality: higher synthetic s′ (sHR=0.64 per 1 cm/s; 95% CI 0.60–0.69; P<0.001), higher synthetic e′ (sHR=0.75 per 1 cm/s; 95% CI 0.71–0.79; P<0.001), and synthetic MAPSE, which demonstrated the largest effect size (sHR=0.15 per 1 cm; 95% CI 0.12–0.19; P<0.001).

After adjustment for age, sex, and conventional ECG abnormalities (Supplemental Table 8), all three synthetic parameters remained independently associated with MI-specific mortality (synthetic s′ adjusted sHR=0.78, 95% CI 0.72–0.84, P<0.001; synthetic e′ adjusted sHR=0.87, 95% CI 0.82–0.93, P<0.001; synthetic MAPSE adjusted sHR=0.49, 95% CI 0.37–0.65, P<0.001).

In a sensitivity analysis using all-cause cardiac mortality as the outcome **(Supplemental Table 9**), all three synthetic parameters remained independently associated with cardiac mortality after adjustment for age, sex, and conventional ECG abnormalities: synthetic s′ (adjusted sHR=0.79, 95% CI 0.76–0.83; P<0.001), synthetic e′ (adjusted sHR=0.91, 95% CI 0.88–0.95; P<0.001), and synthetic MAPSE, which showed the largest effect size (adjusted sHR=0.60, 95% CI 0.51–0.71; P<0.001). Synthetic parameters also added incremental discrimination to a clinical base model in sex- and age-stratified time-dependent AUC analyses (**Supplemental Table 10**).

### Population-cohort prognosis benchmarked with an LV ejection fraction model

In the population cohort, we used the AI-ECG model that predicts incident development of LV systolic dysfunction (EF<40%) over a 1–5 year horizon. Because it was developed to forecast incident heart failure rather than death, the model showed marginal MI-specific mortality discrimination (C-index 0.525; 95% CI 0.507–0.544). Each synthetic TDI parameter added significant incremental discrimination: synthetic MAPSE yielded the largest gain (ΔC=+0.132; 95% CI +0.108 to +0.155; P<0.001), followed by synthetic e′ (ΔC=+0.089; 95% CI +0.066 to +0.112; P<0.001) and synthetic s′ (ΔC=+0.060; 95% CI +0.039 to +0.082; P<0.001) (Supplemental Table 15).

## Discussion

In this study, ECG-derived synthetic LV mechanical parameters were independently associated with adverse outcomes across two complementary clinical settings, and in both they carried prognostic information. In hospitalized AMI patients, synthetic e′ predicted all-cause mortality independently of the GRACE score and, after discharge, independently of both an AI-ECG prediction of reduced ejection fraction and measured echocardiographic LVEF; its adjusted hazard ratio beyond measured LVEF (0.85 per 1 cm/s) was almost identical to that beyond GRACE 2.0 (0.84). In the outpatient population cohort, all three parameters predicted MI-specific mortality with stepwise tertile separation and added discrimination beyond an ECG-based prediction of reduced ejection fraction (ΔC +0.060 to +0.132).

This pattern is consistent with an extensive echocardiographic literature in which longitudinal mechanics [7] — mitral annular velocities [22], longitudinal strain [8,23] and mitral annular plane systolic excursion [24,25] — remain prognostic after adjustment for ejection fraction in both post-infarction and community populations [26], reflecting the subendocardial location of longitudinal fibers and their early involvement in ischemic injury [4,5]. What is new here is the source of that signal: an incremental relationship previously demonstrable only where an echocardiogram had been performed is recovered from the 12-lead ECG alone [12]. These prognostic findings are biologically plausible in light of our recent work in humans with transient LAD occlusion, in which synthetic s′ and e′ deviated early after the onset of ischemia, preceding ST-segment elevation, and recovered more slowly than the ST segment after reperfusion [13]. Revising the classical ischemic cascade in this way supports the idea that the ECG carries an early signal of mechanical dysfunction that conventional ST analysis does not capture, and that diastolic relaxation (e′) is an early and sensitive marker. This is consistent with e′ being the dominant independent predictor in our AMI cohort and with the persistent mechanical dysfunction that can outlast electrical recovery.

These findings build on a growing AI-ECG literature demonstrating that deep learning can identify reduced LVEF [27], detect diastolic dysfunction [12,28], and predict all-cause mortality [12,29] from the surface ECG. However, two limitations have constrained clinical translation of prior work. First, models targeting binary LVEF thresholds may not identify diastolic dysfunction, which develops earlier in the ischemic cascade [30,31]. Second, existing AI-ECG models output scalar probabilities or binary classifications that clinicians cannot examine, interpret, or trend over time. In contrast, synthetic TDI generates continuous velocity and displacement waveforms analogous to conventional echocardiographic longitudinal mechanics, presenting myocardial functional information in a physiologically recognizable format rather than as an isolated prediction score [32]. Importantly, the model was trained with atrial fibrillation-specific augmentation [12], supporting waveform generation in the 7.1% of our cohort with atrial fibrillation on the presenting ECG.

Synthetic TDI parameters remained strongly prognostic even after adjusting for GRACE risk score. Myocardial stunning, microvascular obstruction (affecting up to 50% of STEMI cases), and variable myocyte necrosis each impair longitudinal mechanics [33–35]. This may be particularly relevant in STEMI, where urgent reperfusion priorities frequently preclude comprehensive pre-procedural echocardiography. Serial ECG-derived synthetic assessments obtained at presentation, post-intervention, and discharge could allow longitudinal tracking of functional recovery analogous to serial biomarker assessment [36–38].

The population cohort finding is conceptually distinct. In an unselected outpatient population, Fine-Gray regression appropriately isolates MI-specific risk by accounting for competing deaths [14,15]. A notable difference emerged between cohorts: synthetic e′ was the dominant independent predictor in the AMI cohort, consistent with diastolic relaxation as an early marker of ischemic injury, whereas synthetic MAPSE, an integrated measure of longitudinal displacement, carried the largest incremental discrimination in the stable outpatient cohort, consistent with prior work establishing MAPSE as a marker of cumulative longitudinal dysfunction and adverse remodeling [24,25]. These divergent patterns likely reflect differing biological contexts and warrant further mechanistic study. That the synthetic e′ risk gradient was confined to patients in whom the ECG predicted a preserved ejection fraction reinforces this interpretation, recapitulating a recognized pattern in which diastolic and longitudinal indices retain prognostic value chiefly where systolic function is preserved and ejection fraction is itself poorly discriminating for survival [22,29]. A reduced synthetic e′ in an AMI survivor with a preserved predicted ejection fraction may therefore mark an early, ischemia-defined subset that overlaps mechanistically with the emerging ischemic and coronary microvascular HFpEF phenotype [39], positioning ECG-derived synthetic e′ as a scalable candidate for dedicated study in that group.

Several limitations merit consideration. First, the present analyses establish prognostic association in observational cohorts; the dynamic mechanical fidelity of synthetic TDI during acute ischemia is addressed separately [13] and requires head-to-head comparison against simultaneously measured TDI. Second, because synthetic TDI is derived from the ECG, the prognostic signal may in part reflect a broad ECG-derived cardiovascular risk marker; demonstrating mechanism-specific, incremental value beyond existing ECG-based and clinical markers will require dedicated comparative and prospective study [40], although the synthetic a′ negative control in the population cohort and the ejection-fraction models in the AMI cohort argue against a purely generic ECG risk signal. Third, generalizability across ECG vendors and lead-placement protocols requires further evaluation. Fourth, the median AMI follow-up of 1.44 years may underestimate long-term prognostic contribution. Fifth, registry-based cause-of-death classification is subject to misclassification, particularly with coexisting cardiovascular conditions; a supplemental all-cause cardiac mortality analysis supported robustness. Sixth, the CODE-15 population registry does not capture comorbidity burden, prior cardiovascular disease, or echocardiographic data, precluding direct adjustment for these potential confounders in the population-cohort models; residual confounding by unmeasured comorbidity cannot be excluded. Finally, both cohorts were observational, so prognostic association does not establish that synthetic TDI-guided decisions improve outcomes.

## Conclusions

ECG-derived synthetic TDI parameters were independently associated with mortality in both hospitalized AMI patients and a large outpatient population cohort. These findings support a translational framework in which routinely acquired 12-lead ECGs may provide scalable and physiologically interpretable estimates of LV longitudinal mechanics. Prospective studies are needed to determine the incremental clinical value of synthetic ECG-derived myocardial mechanical assessment in AMI monitoring and cardiovascular risk stratification.

## Data Availability

The clinical cohort data are de-identified electronic health record data from Rutgers Robert Wood Johnson University Hospital and Newark Beth Israel Medical Center. They are not publicly available because of institutional and IRB restrictions. Reasonable requests for de-identified data can be sent to the corresponding author and will be considered, subject to institutional data use agreements and IRB approval. The CODE-15 dataset is publicly available.

## Acknowledgements

This work was supported by the National Heart, Lung, and Blood Institute of the National Institutes of Health (grant R01HL173998). We thank the Telehealth Network of Minas Gerais (TNMG) and the CODE study investigators for providing the CODE-15 cohort data, and the clinical and research staff at Rutgers Robert Wood Johnson University Hospital and Newark Beth Israel Medical Center who supported data collection. The funder had no role in study design; in the collection, analysis, and interpretation of data; in the writing of the report; or in the decision to submit the article for publication. The content is solely the responsibility of the authors and does not necessarily represent the official views of the National Institutes of Health.

## CRediT authorship contribution statement

D.C. and J.F. contributed equally as joint first authors and were responsible for data curation, investigation (electronic health record abstraction and AMI adjudication), formal analysis, and writing – original draft. A.D.J. contributed to data curation, methodology, software, formal analysis, validation, visualization, and writing – original draft and review & editing. S.S. contributed to software and data curation. G.M.M.P. and A.L.P.R. contributed resources, data curation, and investigation for the CODE-15 cohort, and writing – review & editing. J.N., B.R.C. and J.C. contributed to writing – review & editing. N.Y. contributed to conceptualization, methodology, supervision, project administration, and writing – review & editing. P.P.S. conceived and supervised the study, acquired funding (NHLBI R01HL173998), administered the project, interpreted the data, and contributed to writing – original draft and review & editing. D.C., J.F., A.D.J. and S.S. directly accessed and verified the underlying clinical cohort data; G.M.M.P. and A.L.P.R. directly accessed and verified the CODE-15% data. All authors had full access to the study results, critically revised the manuscript for important intellectual content, approved the final version, and accepted responsibility for the decision to submit for publication.

