## Supplemental Material for "ECG-Derived Synthetic Left Ventricular Mechanics Predict Mortality After Acute Myocardial Infarction and Fatal Myocardial Infarction in the General Population"

### Supplemental Methods: Multivariable Logistic Regression for Risk Factor Analysis

The younger non-MI cohort served as a reference for assessing the relationship between biological risk factors and myocardial mechanics, enabling multivariable logistic regression analyses to isolate the independent impact of myocardial infarction on TDI. The MI and non-MI cohorts were pooled and univariate logistic regression models were fit with each risk factor as the binary outcome, including age (>65 years), sex, hypertension, diabetes mellitus, dyslipidemia, obesity, chronic kidney disease, heart failure, reduced LVEF (<50%), and elevated tricuspid regurgitation velocity (TRV >2.8 m/s), and synthetic TDI e′ as the continuous predictor. Multivariable logistic regression models incorporating these covariates were then used to determine whether synthetic TDI parameters independently differentiated infarcted from non-infarcted individuals. Results are expressed as odds ratios (ORs) with 95% confidence intervals. Parallel models were fit using corresponding measured TDI parameters to directly compare the direction and magnitude of associations between synthetic and measured waveforms.

### Supplemental Methods: Incremental Prognostic Value Beyond GRACE 2.0 and Beyond Ejection Fraction

To evaluate incremental prognostic value above established clinical risk stratification, nested Cox models were fit with GRACE 2.0 alone as the reference and with each synthetic parameter added individually. Model comparisons used likelihood-ratio (LR) $\chi^2$ tests with degrees of freedom equal to the number of added covariates. This comparison was performed in two settings: (i) the full, all-comers AMI cohort; and (ii) the post-discharge cohort (N=1,010 with a complete GRACE 2.0 score; 61 deaths over the full post-discharge follow-up window), analyzed both as continuous synthetic parameters (exploratory) and by categorical e′ tertile.

Incremental value beyond ejection fraction was assessed in the same post-discharge cohort. Ejection fraction was represented in four ways: AI-ECG predictions of a concurrent LVEF below 50% from AiTiA and from HeartWise; the AI-ECG HFrEF risk model of Pan et al., which predicts incident LVEF of 40% or less over a 1 to 5 year horizon; and measured echocardiographic LVEF below 50% as the reference standard. The three AI-ECG models were dichotomized at a predicted probability of 0.5. For each representation, a Cox model containing the ejection-fraction term alone was compared with models adding one synthetic index at a time (synthetic e′, s′, MAPSE and a′), each entered per 1 cm/s or, for MAPSE, per 1 cm. The likelihood-ratio chi-square attributable to the synthetic index (1 degree of freedom) is reported both in absolute terms and as a percentage of the full model chi-square, alongside the

Harrell C-index and the change in C-index relative to the ejection-fraction term alone. Synthetic a′ was pre-specified as a negative control, since late diastolic (atrial) annular velocity should carry little ventricular prognostic information. Results are reported in Supplemental Table 12. The prognostic value of each ejection-fraction representation on its own, before any synthetic index is added, is reported separately in Supplemental Table 11. Synthetic e′ was additionally entered as two tertile indicators with the lowest tertile as reference, using the full clinical AMI cohort boundaries, giving a 2 degree of freedom likelihood-ratio test within each ejection-fraction representation; those models are in Supplemental Table 14, and the corresponding two-year Kaplan-Meier mortality by predicted ejection fraction and e′ tertile, which is the analysis plotted in Figure 4 Panel B, is tabulated in Supplemental Table 13.

In the population cohort, incremental discrimination beyond the AI-ECG HFrEF risk model was quantified with the Wolbers competing-risk concordance index for MI-specific mortality, in which competing (non-MI) deaths are treated as non-events rather than as censored observations. Each synthetic index was added singly to the dichotomized comparator, and synthetic s′ and MAPSE were also added together. Confidence intervals for the change in concordance were derived from paired concordance influence functions, so that every comparison is made within the same participants. Results are reported in Supplemental Table 15.

**Supplemental Table 1.** Baseline Electrocardiographic and Synthetic Echocardiographic Characteristics of the Study Cohorts

| Variable | Clinical, (N=2,011) | Non-MI (N=946) | MI (N=1,065) | Population (N=219,564) |
|---|---|---|---|---|
| **Demographics and clinical characteristics** | | | | |
| Age >65 years, n (%) | 823 (40.9) | 307 (32.5) | 516 (48.5) | - |
| Male sex, n (%) | 1,205 (59.9) | 481 (50.8) | 724 (68.0) | 86,977 (39.6) |
| Hypertension, n (%) | 1,233 (61.3) | 459 (48.5) | 774 (72.7) | - |
| Diabetes mellitus, n (%) | 641 (31.9) | 191 (20.2) | 450 (42.3) | - |
| Dyslipidemia, n (%) | 796 (39.6) | 255 (27.0) | 541 (50.8) | - |
| Obesity, n (%) | 243 (12.1) | 89 (9.4) | 154 (14.5) | - |
| Chronic kidney disease, n (%) | 291 (14.5) | 70 (7.4) | 221 (20.8) | - |
| Heart failure, n (%) | 335 (16.7) | 82 (8.7) | 253 (23.8) | - |
| Reduced LVEF (<50%), n (%) | 525 (26.5) | 100 (10.6) | 425 (40.9) | - |
| Elevated TRV (>2.8 m/s), n (%) | 189 (18.1) | 93 (15.1) | 96 (22.6) | - |
| **ECG findings, n (%)** | | | | |
| 1st-degree AV block | 18 (0.9) | 9 (1.0) | 9 (0.8) | 2,943 (1.3) |
| Right bundle branch block | 234 (11.6) | 92 (9.7) | 142 (13.3) | 5,041 (2.3) |
| Left bundle branch block | 105 (5.2) | 43 (4.6) | 62 (5.8) | 3,083 (1.4) |
| Sinus bradycardia | 257 (12.8) | 151 (16.0) | 106 (10.0) | 1,456 (0.7) |
| ST-T abnormalities | 584 (29.0) | 168 (17.8) | 416 (39.1) | 5,088 (2.3) |
| Atrial fibrillation on the index ECG | 131 (6.5) | 55 (5.8) | 76 (7.1) | 3,149 (1.4) |
| **Synthetic echocardiographic variables, median (IQR)** | | | | |
| Synthetic s′ (avg), cm/s | 6.4 (5.7–6.9) | 6.7 (6.1–7.0) | 6.1 (5.3–6.7) | 6.1 (6.0–6.3) |
| Synthetic e′ (avg), cm/s | 7.8 (5.9–9.7) | 8.7 (6.8–10.4) | 7.2 (5.4–9.0) | 5.5 (4.5–6.3) |
| Synthetic MAPSE (avg), cm | 1.7 (1.3–2.0) | 1.8 (1.5–2.1) | 1.6 (1.1–1.9) | 1.1 (0.9–1.3) |

Values are median (interquartile range) for continuous variables and n (%) for categorical variables. Atrial fibrillation in this table denotes atrial fibrillation present on the index 12-lead ECG; a documented history of atrial fibrillation at any time prior to or during the index admission is reported separately in Table 1. Percentages for reduced LVEF and for elevated tricuspid regurgitation velocity are calculated against the patients in whom that measurement was available, not against the full cohort; tricuspid regurgitation velocity in particular was unrecorded in 640 of 1,065 AMI patients and 329 of 946 controls. Abbreviations: AV, atrioventricular; e′, early diastolic mitral annular velocity; LVEF, left ventricular ejection fraction; MAPSE, mitral annular plane systolic excursion; MI, myocardial infarction; s′, peak systolic mitral annular velocity; TRV, tricuspid regurgitation velocity.

**Supplemental Table 2.** Univariable and Multivariable Logistic Regression for AMI Status: Measured vs. Synthetic Early Diastolic Velocity (e′), Adjusted for Cardiovascular Risk Factors.

| Predictor | Univariate OR (95% CI) | P value | Multivariate OR (95% CI) | P value |
|---|---|---|---|---|
| **Model 1: Measured e′ and Clinical Risk Factors** | | | | |
| Measured e′ (per 1 unit increase) | 0.74 (0.71–0.77) | <0.001 | 0.76 (0.72–0.79) | <0.001 |
| Age >65 years | 1.81 (1.50–2.19) | <0.001 | 0.87 (0.69–1.10) | 0.248 |
| Hypertension | 2.73 (2.25–3.32) | <0.001 | 1.10 (0.85–1.41) | 0.477 |
| Diabetes mellitus | 2.78 (2.26–3.42) | <0.001 | 1.64 (1.28–2.09) | <0.001 |
| Smoking | 11.41 (7.31–18.83) | <0.001 | 12.24 (7.64–20.67) | <0.001 |
| Obesity | 1.54 (1.16–2.05) | 0.003 | 1.06 (0.76–1.48) | 0.737 |
| Dyslipidemia | 2.74 (2.26–3.33) | <0.001 | 1.68 (1.33–2.12) | <0.001 |
| Chronic kidney disease | 3.02 (2.26–4.06) | <0.001 | 1.88 (1.35–2.63) | <0.001 |
| **Model 2: Synthetic e′ and Clinical Risk Factors** | | | | |
| Synthetic e′ (per 1 unit increase) | 0.81 (0.78–0.84) | <0.001 | 0.85 (0.81–0.88) | <0.001 |
| Age >65 years | 1.96 (1.63–2.35) | <0.001 | 1.21 (0.97–1.50) | 0.084 |
| Hypertension | 2.82 (2.35–3.40) | <0.001 | 1.32 (1.04–1.68) | 0.020 |
| Diabetes mellitus | 2.89 (2.37–3.54) | <0.001 | 1.85 (1.47–2.33) | <0.001 |
| Smoking | 11.51 (7.39–18.94) | <0.001 | 12.89 (8.13–21.54) | <0.001 |
| Obesity | 1.63 (1.24–2.15) | <0.001 | 1.09 (0.80–1.50) | 0.575 |
| Dyslipidemia | 2.80 (2.32–3.38) | <0.001 | 1.70 (1.37–2.13) | <0.001 |
| Chronic kidney disease | 3.28 (2.48–4.38) | <0.001 | 1.97 (1.44–2.72) | <0.001 |

Sample size: N=2,011 (total pooled MI/non-MI cohort); n=1,849 complete cases in the measured e′ models and n=2,011 in the synthetic e′ models. Outcome: MI vs. non-MI status; reference group: non-MI. Both measured and synthetic e′ remained independently and significantly associated with AMI status after adjustment for age, hypertension, diabetes mellitus, smoking, obesity, dyslipidemia, and chronic kidney disease. Abbreviations: OR, odds ratio; CI, confidence interval. The two blocks therefore rest on different denominators: measured e′ is missing in 125 AMI patients and 37 non-AMI controls, so the measured-e′ models are fit on 1,849 rows and the synthetic-e′ models on 2,011. Confidence intervals are profile-likelihood intervals, used as the primary estimator throughout this table.

**Supplemental Table 3.** Synthetic TDI Tertile Boundaries by Cohort.

| Cohort | Parameter | Low Tertile | Mid Tertile | High Tertile |
|---|---|---|---|---|
| **Clinical Cohort (full AMI, N=1,065)** | Synthetic s′ | ≤5.69 cm/s | >5.69 to ≤6.56 cm/s | >6.56 cm/s |
| | Synthetic e′ | ≤5.89 cm/s | >5.89 to ≤8.33 cm/s | >8.33 cm/s |
| | Synthetic a′ | ≤4.82 cm/s | >4.82 to ≤5.95 cm/s | >5.95 cm/s |
| | Synthetic MAPSE | ≤1.32 cm | >1.32 to ≤1.76 cm | >1.76 cm |
| **Population Cohort (N=219,564)** | Synthetic s′ | ≤6.06 cm/s | >6.06 to ≤6.24 cm/s | >6.24 cm/s |
| | Synthetic e′ | ≤4.87 cm/s | >4.87 to ≤6.00 cm/s | >6.00 cm/s |
| | Synthetic a′ | ≤5.72 cm/s | >5.72 to ≤6.18 cm/s | >6.18 cm/s |
| | Synthetic MAPSE | ≤0.96 cm | >0.96 to ≤1.25 cm | >1.25 cm |

Clinical-cohort boundaries were derived in the full AMI cohort (N=1,065) and applied unchanged wherever tertiles are used, including the post-discharge cohort in Supplemental Tables 5 and 6 and in Figure 4 Panel B. Population-cohort boundaries were derived within the population cohort and applied unchanged. Abbreviations: a′, late diastolic mitral annular velocity; e′, early diastolic mitral annular velocity; MAPSE, mitral annular plane systolic excursion; s′, peak systolic mitral annular velocity; TDI, tissue Doppler imaging.

**Supplemental Table 4.** Univariate and Covariate-Adjusted Cox Proportional-Hazards Models for All-Cause Mortality by Synthetic Parameter (Clinical AMI Cohort).

| Covariate | Univariate HR (95% CI) | P | Model 1: Synthetic s′ HR (95% CI) | P | Model 2: Synthetic e′ HR (95% CI) | P | Model 3: Synthetic MAPSE HR (95% CI) | P |
|---|---|---|---|---|---|---|---|---|
| **Synthetic Parameters** | | | | | | | | |
| Synthetic s′ (per 1 cm/s) | 0.80 (0.71–0.92) | 0.001 | 0.90 (0.78–1.04) | 0.143 | - | - | - | - |
| Synthetic e′ (per 1 cm/s) | 0.85 (0.79–0.92) | <0.001 | - | - | 0.90 (0.84–0.98) | 0.012 | - | - |
| Synthetic MAPSE (per 1 cm) | 0.57 (0.41–0.80) | 0.001 | - | - | - | - | 0.77 (0.54–1.09) | 0.144 |
| **Clinical Covariates (Retained if Significant on Univariable Analysis)** | | | | | | | | |
| Age (per year) | 1.05 (1.04–1.07) | <0.001 | 1.04 (1.02–1.06) | <0.001 | 1.04 (1.02–1.06) | <0.001 | 1.04 (1.02–1.06) | <0.001 |
| Chronic kidney disease | 2.14 (1.44–3.19) | <0.001 | 1.37 (0.90–2.09) | 0.146 | 1.36 (0.89–2.07) | 0.154 | 1.37 (0.90–2.10) | 0.141 |
| Atrial fibrillation | 2.23 (1.47–3.36) | <0.001 | 1.25 (0.80–1.95) | 0.336 | 1.21 (0.77–1.89) | 0.411 | 1.24 (0.79–1.94) | 0.351 |
| Prior cardiac arrest | 3.11 (1.90–5.11) | <0.001 | 2.47 (1.50–4.08) | <0.001 | 2.56 (1.55–4.23) | <0.001 | 2.50 (1.51–4.12) | <0.001 |
| **Coronary Territory (Ref: Non-Localized/Other)** | | | | | | | | |
| Septal (LAD) | 1.43 (0.93–2.21) | 0.106 | 1.31 (0.84–2.03) | 0.228 | 1.26 (0.81–1.96) | 0.297 | 1.27 (0.81–1.98) | 0.300 |
| Lateral (LCx/Diagonal) | 1.66 (0.94–2.95) | 0.083 | 1.49 (0.84–2.65) | 0.171 | 1.50 (0.85–2.67) | 0.165 | 1.46 (0.82–2.59) | 0.200 |
| Inferior (RCA/PDA) | 1.01 (0.48–2.13) | 0.972 | 1.25 (0.59–2.64) | 0.561 | 1.25 (0.59–2.64) | 0.557 | 1.22 (0.58–2.58) | 0.602 |

Models 1–3 each evaluate one synthetic parameter separately, adjusted for baseline regional ECG-defined coronary vascular territory (septal [LAD], lateral [LCx/diagonal], inferior [RCA/PDA], with non-localized/other as reference) and clinical covariates that were significantly associated with all-cause mortality on univariable analysis from a candidate list (age, sex, hypertension, diabetes mellitus, smoking, obesity, dyslipidemia, chronic kidney disease, atrial fibrillation, coronary territory, and prior cardiac arrest); sex, hypertension, diabetes mellitus, smoking, obesity, and dyslipidemia were not univariably associated with mortality (all P>0.05) and were not retained in the final models. All covariates included in these models were complete across the study cohort (N=1,065). Abbreviations: CI, confidence interval; HR, hazard ratio; LAD, left anterior descending artery; LCx, left circumflex artery; MAPSE, mitral annular plane systolic excursion; PDA, posterior descending artery; RCA, right coronary artery; s′, synthetic peak systolic mitral annular velocity; e′, synthetic early diastolic mitral annular velocity.

**Supplemental Table 5.** GRACE and Single Synthetic Parameters: Complete and Post-Discharge AMI Cohorts.

| Model | Univariable HR (95% CI), P | GRACE 2.0-adjusted HR (95% CI), P | C-index (95% CI) | LR P vs GRACE 2.0 |
|---|---|---|---|---|
| **Complete AMI all-comer cohort (N=1,058 with a complete GRACE 2.0 score; 109 deaths)** | | | | |
| GRACE 2.0 alone | 1.23 (1.18–1.28), P<0.001 | -* | 0.78 (0.74–0.82) | - |
| GRACE 2.0 + synthetic s′ | 0.81 (0.71–0.92), P=0.001 | 0.90 (0.78–1.03), P=0.117 | 0.78 (0.74–0.82) | 0.125 |
| GRACE 2.0 + synthetic e′ | 0.85 (0.79–0.92), P<0.001 | 0.90 (0.83–0.97), P=0.006 | 0.79 (0.75–0.83) | 0.0058 |
| GRACE 2.0 + synthetic a′ | 0.99 (0.87–1.12), P=0.855 | 1.03 (0.92–1.15), P=0.611 | 0.78 (0.74–0.82) | 0.611 |
| GRACE 2.0 + synthetic MAPSE | 0.57 (0.41–0.80), P=0.001 | 0.79 (0.57–1.10), P=0.157 | 0.78 (0.74–0.82) | 0.16 |
| **Post-discharge AMI cohort, full post-discharge follow-up (N=1,010 with a complete GRACE 2.0 score; 61 deaths)** | | | | |
| GRACE 2.0 alone | 1.14 (1.08–1.21), P<0.001 | -* | 0.67 (0.60–0.74) | - |
| GRACE 2.0 + synthetic s′ | 0.77 (0.65–0.91), P=0.002 | 0.82 (0.69–0.97), P=0.020 | 0.68 (0.61–0.75) | 0.027 |
| GRACE 2.0 + synthetic e′ | 0.82 (0.74–0.90), P<0.001 | 0.84 (0.76–0.93), P=0.001 | 0.70 (0.64–0.77) | <0.001 |
| GRACE 2.0 + synthetic a′ | 1.01 (0.86–1.20), P=0.862 | 1.03 (0.88–1.21), P=0.699 | 0.67 (0.60–0.74) | 0.700 |
| GRACE 2.0 + synthetic MAPSE | 0.49 (0.31–0.76), P=0.001 | 0.59 (0.38–0.92), P=0.021 | 0.68 (0.61–0.75) | 0.023 |

Hazard ratios for the synthetic indices are per 1 cm/s, or per 1 cm for MAPSE; lower e′, lower s′ and lower MAPSE all indicate worse left ventricular long-axis mechanics. The GRACE 2.0 alone rows report the hazard ratio per 10 GRACE 2.0 points, not per 1 point; these models are unadjusted, so no separate GRACE 2.0-adjusted estimate is given. *GRACE 2.0 remained significantly associated with mortality in every combined model (all P<0.001), with its hazard ratio per 10 points changing by at most 0.6% in the complete AMI cohort and 1.3% in the post-discharge cohort; the largest attenuation in both cohorts occurred with synthetic e′, and none occurred with the synthetic a′ negative control. Univariable C-indices in the complete AMI cohort were 0.60 for e′, 0.60 for s′ and 0.59 for MAPSE. Likelihood-ratio tests compare GRACE 2.0 plus one synthetic index against GRACE 2.0 alone (df=1). The post-discharge cohort is analyzed over the full post-discharge follow-up window. Synthetic a′ is included as a negative control. Abbreviations: CI, confidence interval; df, degrees of freedom; GRACE 2.0, updated Global Registry of Acute Coronary Events risk model; HR, hazard ratio; LR, likelihood ratio; MAPSE, mitral annular plane systolic excursion.

**Supplemental Table 6.** Post-Discharge All-Cause Mortality by Synthetic e′ Tertile, GRACE 2.0-Adjusted (Post-Discharge Cohort, N=1,010, Full Follow-Up)

| Synthetic e′ Tertile | Tertile Cut-Point | Post-Discharge Mortality, n/N (%) | GRACE-Adjusted HR (95% CI) | P value |
|---|---|---|---|---|
| Lowest (T1) | ≤5.89 cm/s | 31/335 (9.3%) | 2.72 (1.32–5.60) | 0.006 |
| Middle (T2) | >5.89 to ≤8.33 cm/s | 20/334 (6.0%) | 1.81 (0.85–3.89) | 0.126 |
| Highest (T3) | >8.33 cm/s | 10/341 (2.9%) | Reference | - |

Tertile cut points were derived in the full clinical AMI cohort (Supplemental Table 3) and applied unchanged to this post-discharge cohort, the same population analyzed in the post-discharge section of Supplemental Table 5. Hazard ratios are adjusted for GRACE 2.0 and reference the highest tertile. The two-degree-of-freedom likelihood-ratio test for the tertile term gives P = 0.014. Abbreviations: CI, confidence interval; GRACE 2.0, updated Global Registry of Acute Coronary Events risk model; HR, hazard ratio.

**Supplemental Table 7.** Baseline Characteristics of the Population Cohort by Vital Status and Adjudicated Cause of Death

| Characteristic | Overall | Alive | Dead | P-value |
|---|---|---|---|---|
| N | 219,564 | 208,829 | 10,735 | - |
| **Demographics** | | | | |
| Age, years, Median (IQR) | 50 (33–67) | 49 (33–66) | 75 (64–81) | <0.001 |
| Male sex, n (%) | 86,977 (39.6) | 81,280 (38.9) | 5,697 (53.1) | <0.001 |
| **ECG Findings, n (%)** | | | | |
| Normal ECG | 89,514 (40.8) | 87,746 (42.0) | 1,768 (16.5) | <0.001 |
| 1st-degree AV block | 2,943 (1.3) | 2,595 (1.2) | 348 (3.2) | <0.001 |
| Right bundle branch block | 5,041 (2.3) | 4,403 (2.1) | 638 (5.9) | <0.001 |
| Left bundle branch block | 3,083 (1.4) | 2,598 (1.2) | 485 (4.5) | <0.001 |
| Sinus bradycardia | 1,456 (0.7) | 1,409 (0.7) | 47 (0.4) | 0.004 |
| ST-T abnormalities | 5,088 (2.3) | 4,592 (2.2) | 496 (4.6) | <0.001 |
| Atrial fibrillation | 3,149 (1.4) | 2,365 (1.1) | 784 (7.3) | <0.001 |
| **Synthetic Echocardiographic Parameters, Median (IQR)** | | | | |
| Synthetic s′ (avg), cm/s | 6.15 (5.99–6.29) | 6.15 (6.00–6.29) | 6.10 (5.82–6.27) | <0.001 |
| Synthetic e′ (avg), cm/s | 5.45 (4.54–6.34) | 5.47 (4.55–6.36) | 5.05 (4.28–5.95) | <0.001 |
| Synthetic MAPSE (avg), cm | 1.09 (0.90–1.35) | 1.09 (0.90–1.36) | 0.97 (0.81–1.18) | <0.001 |
| **Adjudicated Cause of Death, n (%)** | | | | |
| Cardiac death | 2,566 (1.2) | - | 2,566 (23.9) | - |
| Heart failure death | 887 (0.4) | - | 887 (8.3) | - |
| Myocardial infarction death | 817 (0.4) | - | 817 (7.6) | - |
| Other cardiac death | 862 (0.4) | - | 862 (8.0) | - |
| Non-cardiac death | 8,169 (3.7) | - | 8,169 (76.1) | - |

Values are presented as median (interquartile range) or n (%). P-values from Mann-Whitney U test for continuous variables and chi-squared test for categorical variables. This subset includes all patients with adjudicated cause-specific mortality data. Non-cardiac death column percentage (3.7%) reflects share of the overall cohort; row percentage among deaths (76.1%) shown in the Dead column. Abbreviations: AV, atrioventricular; ECG, electrocardiogram; MAPSE, mitral annular plane systolic excursion. Atrial fibrillation here denotes atrial fibrillation present on the index 12-lead ECG.

**Supplemental Table 8.** Univariable and Covariate-Adjusted Fine–Gray Subdistribution Hazard Models for MI-Specific Mortality (Population Cohort)

| Covariate | Univariable sHR (95% CI) | P | Model 1: Synthetic s′ | P | Model 2: Synthetic e′ | P | Model 3: Synthetic MAPSE | P |
|---|---|---|---|---|---|---|---|---|
| **Synthetic parameters (each modelled separately)** | | | | | | | | |
| Synthetic s′ (per 1 cm/s) | 0.64 (0.60–0.69) | <0.001 | 0.78 (0.72–0.84) | <0.001 | - | - | - | - |
| Synthetic e′ (per 1 cm/s) | 0.75 (0.71–0.79) | <0.001 | - | - | 0.87 (0.82–0.93) | <0.001 | - | - |
| Synthetic MAPSE (per 1 cm) | 0.15 (0.12–0.19) | <0.001 | - | - | - | - | 0.49 (0.37–0.65) | <0.001 |
| Synthetic a′ (per 1 cm/s) | 0.87 (0.78–0.96) | 0.006 | - | - | - | - | - | - |
| **Clinical covariates** | | | | | | | | |
| Age (per year) | 1.07 (1.06–1.07) | <0.001 | 1.06 (1.06–1.07) | <0.001 | 1.06 (1.06–1.07) | <0.001 | 1.06 (1.06–1.07) | <0.001 |
| Male sex | 2.21 (1.92–2.54) | <0.001 | 2.04 (1.77–2.35) | <0.001 | 2.13 (1.85–2.46) | <0.001 | 2.11 (1.83–2.44) | <0.001 |
| Right bundle branch block | 2.13 (1.56–2.90) | <0.001 | 1.06 (0.78–1.45) | 0.713 | 1.08 (0.79–1.47) | 0.639 | 0.99 (0.73–1.36) | 0.974 |
| Left bundle branch block | 4.77 (3.62–6.28) | <0.001 | 2.00 (1.50–2.67) | <0.001 | 2.15 (1.62–2.85) | <0.001 | 1.85 (1.38–2.48) | <0.001 |
| ST-T abnormality | 1.74 (1.22–2.47) | 0.002 | 1.74 (1.22–2.49) | 0.002 | 1.75 (1.23–2.50) | 0.002 | 1.84 (1.29–2.62) | <0.001 |
| Atrial fibrillation | 4.34 (3.27–5.76) | <0.001 | 1.54 (1.16–2.05) | 0.003 | 1.57 (1.18–2.09) | 0.002 | 1.60 (1.20–2.12) | 0.001 |
| 1st-degree AV block | 2.76 (1.93–3.95) | <0.001 | 1.23 (0.85–1.77) | 0.266 | 1.20 (0.84–1.74) | 0.318 | 1.20 (0.84–1.73) | 0.320 |

Models 1 to 3 each contain one synthetic parameter together with age, sex, right bundle branch block, left bundle branch block, ST-T abnormality, atrial fibrillation and first-degree atrioventricular block; non-MI death is the competing event. Subdistribution hazard ratios are per 1 cm/s, or per 1 cm for MAPSE. Synthetic a′ is shown univariably as a negative control. Models were fitted by risk-set-weighted Cox regression (survival::finegray), which reproduces cmprsk::crr to five decimal places in this cohort and scales to 219,564 observations. Abbreviations: AV, atrioventricular; CI, confidence interval; MAPSE, mitral annular plane systolic excursion; sHR, subdistribution hazard ratio.

**Supplemental Table 9.** Sensitivity Analysis: All-Cause Cardiac Mortality as Outcome (Population Cohort)

| Covariate | Univariable sHR (95% CI) | P | Model 1: Synthetic s′ | P | Model 2: Synthetic e′ | P | Model 3: Synthetic MAPSE | P |
|---|---|---|---|---|---|---|---|---|
| **Synthetic parameters (each modelled separately)** | | | | | | | | |
| Synthetic s′ (per 1 cm/s) | 0.64 (0.61–0.66) | <0.001 | 0.79 (0.76–0.83) | <0.001 | - | - | - | - |
| Synthetic e′ (per 1 cm/s) | 0.76 (0.74–0.78) | <0.001 | - | - | 0.91 (0.88–0.95) | <0.001 | - | - |
| Synthetic MAPSE (per 1 cm) | 0.15 (0.13–0.18) | <0.001 | - | - | - | - | 0.60 (0.51–0.71) | <0.001 |
| Synthetic a′ (per 1 cm/s) | 0.83 (0.78–0.87) | <0.001 | - | - | - | - | - | - |
| **Clinical covariates** | | | | | | | | |
| Age (per year) | 1.07 (1.07–1.07) | <0.001 | 1.06 (1.06–1.07) | <0.001 | 1.06 (1.06–1.07) | <0.001 | 1.06 (1.06–1.06) | <0.001 |
| Male sex | 1.69 (1.57–1.83) | <0.001 | 1.50 (1.39–1.62) | <0.001 | 1.54 (1.42–1.67) | <0.001 | 1.54 (1.42–1.67) | <0.001 |
| Right bundle branch block | 3.52 (3.06–4.05) | <0.001 | 1.72 (1.48–1.99) | <0.001 | 1.75 (1.51–2.02) | <0.001 | 1.64 (1.42–1.90) | <0.001 |
| Left bundle branch block | 5.49 (4.73–6.36) | <0.001 | 2.24 (1.92–2.62) | <0.001 | 2.41 (2.06–2.81) | <0.001 | 2.17 (1.85–2.54) | <0.001 |
| ST-T abnormality | 1.74 (1.42–2.12) | <0.001 | 1.73 (1.41–2.12) | <0.001 | 1.75 (1.43–2.15) | <0.001 | 1.80 (1.47–2.21) | <0.001 |
| Atrial fibrillation | 9.25 (8.20–10.43) | <0.001 | 3.34 (2.94–3.79) | <0.001 | 3.44 (3.03–3.90) | <0.001 | 3.47 (3.06–3.93) | <0.001 |
| 1st-degree AV block | 3.22 (2.66–3.89) | <0.001 | 1.45 (1.19–1.76) | <0.001 | 1.42 (1.17–1.73) | <0.001 | 1.42 (1.17–1.73) | <0.001 |

Model structure is as in Supplemental Table 8, with non-cardiac death as the competing event. Adjusted subdistribution hazard ratios for the synthetic indices are 0.79 for s′, 0.91 for e′ and 0.60 for MAPSE. Findings are directionally concordant with the MI-specific mortality analysis in Supplemental Table 8. Abbreviations: AV, atrioventricular; CI, confidence interval; MAPSE, mitral annular plane systolic excursion; sHR, subdistribution hazard ratio.

**Supplemental Table 10.** Incremental Benefit of Synthetic LV Mechanics on Time-Dependent AUC for MI Mortality, Stratified by Sex and Age (Population Cohort)

| Time point | Base AUC (95% CI) | Base + synthetic TDI AUC (95% CI) | Delta AUC (95% CI) |
|---|---|---|---|
| **Male, all ages (n = 86,977; 481 MI deaths)** | | | |
| 1 year | 0.793 (0.767–0.820) | 0.805 (0.778–0.831) | +0.013 (+0.002 to +0.023) |
| 3 years | 0.775 (0.755–0.793) | 0.791 (0.772–0.810) | +0.016 (+0.009 to +0.023) |
| 5 years | 0.764 (0.743–0.786) | 0.781 (0.762–0.802) | +0.017 (+0.008 to +0.026) |
| **Female, all ages (n = 132,587; 336 MI deaths)** | | | |
| 1 year | 0.829 (0.793–0.862) | 0.830 (0.795–0.862) | +0.001 (–0.005 to +0.006) |
| 3 years | 0.821 (0.797–0.842) | 0.824 (0.800–0.844) | +0.002 (–0.002 to +0.007) |
| 5 years | 0.799 (0.775–0.822) | 0.803 (0.778–0.825) | +0.003 (–0.000 to +0.007) |
| **Age < 75 years, both sexes (n = 186,779; 434 MI deaths)** | | | |
| 1 year | 0.627 (0.586–0.668) | 0.714 (0.665–0.761) | +0.086 (+0.050 to +0.122) |
| 3 years | 0.621 (0.593–0.650) | 0.714 (0.683–0.748) | +0.093 (+0.070 to +0.120) |
| 5 years | 0.623 (0.596–0.650) | 0.725 (0.696–0.752) | +0.102 (+0.079 to +0.126) |
| **Age ≥ 75 years, both sexes (n = 32,785; 383 MI deaths)** | | | |
| 1 year | 0.567 (0.526–0.607) | 0.611 (0.557–0.660) | +0.044 (+0.013 to +0.073) |
| 3 years | 0.574 (0.542–0.602) | 0.610 (0.568–0.646) | +0.036 (+0.015 to +0.057) |
| 5 years | 0.572 (0.545–0.602) | 0.604 (0.567–0.636) | +0.031 (+0.013 to +0.049) |

The base model contains age in the sex-stratified analysis and sex in the age-stratified analysis; the augmented model adds synthetic e′, s′ and MAPSE. Time-dependent AUCs are inverse-probability-of-censoring weighted under the Fine-Gray competing-risks framework, with non-MI death as the competing event. Horizons stop at 5 years: the censoring survival function, estimated by the reverse Kaplan-Meier of the censoring distribution, is 0.96, 0.60 and 0.28 at 1, 3 and 5 years but falls to 0.040 at 7 years and 0.00013 at 10 years, so the censoring weights are undefined beyond 5 years. Confidence intervals come from a paired bootstrap with 400 replicates. P values are not reported for this table. The smallest value attainable at 400 replicates is 0.005, and eight of the twelve comparisons sat exactly at that floor, so a P value here would describe the resampling budget rather than the strength of the evidence; the confidence interval for the AUC difference carries the inference. Abbreviations: AUC, area under the receiver operating characteristic curve; CI, confidence interval; MI, myocardial infarction; TDI, tissue Doppler imaging.

### Supplemental Table 11. Association of Each Dichotomized Ejection-Fraction Prediction With All-Cause Death (Post-Discharge AMI Cohort)

| Prediction | N predicted reduced | Deaths, predicted reduced | Deaths, predicted preserved | HR (95% CI) | P |
|---|---|---|---|---|---|
| AiTiA — predicted LVEF < 50% | 159 | 22 / 159 (13.8%) | 38 / 857 (4.4%) | 3.94 (2.32–6.67) | < 0.001 |
| AI-ECG HFrEF risk — predicted LVEF ≤ 40% | 162 | 19 / 162 (11.7%) | 42 / 855 (4.9%) | 2.89 (1.68–4.99) | < 0.001 |
| HeartWise — predicted LVEF < 50% | 162 | 13 / 162 (8.0%) | 48 / 855 (5.6%) | 1.47 (0.80–2.71) | 0.218 |
| Measured echo LVEF < 50% | 402 | 35 / 402 (8.7%) | 24 / 595 (4.0%) | 2.50 (1.49–4.21) | < 0.001 |

Each row is a Cox model containing that ejection-fraction term alone, fit in the post-discharge AMI cohort. All three AI-ECG models were dichotomized at a predicted probability of 0.5. AiTiA and HeartWise predict a concurrent LVEF below 50%; the AI-ECG HFrEF risk model of Pan et al. predicts incident heart failure with reduced ejection fraction, LVEF 40% or below. Measured echocardiographic LVEF below 50% flags 402 of 997 patients, roughly 2.5 times as many as the AI-ECG tests at a 0.5 threshold. Denominators differ across rows because AiTiA is missing in one patient and measured LVEF in twenty.

**Supplemental Table 12. Synthetic Tissue Doppler Indices Added to Each Representation of Ejection Fraction (Post-Discharge AMI Cohort)**

| Index | Adjusted HR (95% CI) | P | LR chi-square (1 df) | % of model chi-square | C-index | Delta C vs EF alone |
|---|---|---|---|---|---|---|
| **Added to AiTiA, predicted LVEF < 50% (n=1,016; 60 deaths; EF term HR 3.94, 95% CI 2.32–6.67, P<0.001)** | | | | | | |
| Synthetic e′ (per 1 cm/s) | 0.86 (0.77–0.95) | 0.004 | 8.69 | 28% | 0.692 | +0.079 |
| Synthetic s′ (per 1 cm/s) | 0.83 (0.70–1.00) | 0.047 | 3.64 | 14% | 0.678 | +0.065 |
| Synthetic MAPSE (per 1 cm) | 0.62 (0.39–1.00) | 0.048 | 3.81 | 15% | 0.677 | +0.064 |
| Synthetic a′ (per 1 cm/s) | 1.07 (0.91–1.26) | 0.423 | 0.64 | 3% | 0.630 | +0.017 |
| **Added to AI-ECG HFrEF risk (LVEF ≤ 40%) of Pan et al. (n=1,017; 61 deaths; EF term HR 2.89, 95% CI 1.68–4.99, P<0.001)** | | | | | | |
| Synthetic e′ (per 1 cm/s) | 0.84 (0.76–0.93) | 0.001 | 10.62 | 46% | 0.685 | +0.089 |
| Synthetic s′ (per 1 cm/s) | 0.83 (0.69–0.99) | 0.042 | 3.77 | 23% | 0.662 | +0.066 |
| Synthetic MAPSE (per 1 cm) | 0.60 (0.37–0.96) | 0.032 | 4.43 | 26% | 0.663 | +0.068 |
| Synthetic a′ (per 1 cm/s) | 1.05 (0.90–1.23) | 0.518 | 0.41 | 3% | 0.624 | +0.028 |
| **Added to HeartWise, predicted LVEF < 50% (n=1,017; 61 deaths; EF term HR 1.47, 95% CI 0.80–2.71, P=0.218)** | | | | | | |
| Synthetic e′ (per 1 cm/s) | 0.82 (0.74–0.91) | <0.001 | 15.36 | 92% | 0.644 | +0.111 |
| Synthetic s′ (per 1 cm/s) | 0.77 (0.66–0.91) | 0.003 | 7.97 | 85% | 0.629 | +0.096 |
| Synthetic MAPSE (per 1 cm) | 0.50 (0.32–0.78) | 0.002 | 8.84 | 86% | 0.621 | +0.088 |
| Synthetic a′ (per 1 cm/s) | 1.02 (0.86–1.20) | 0.836 | 0.04 | 3% | 0.547 | +0.014 |
| **Added to measured echocardiographic LVEF < 50% (n=997; 59 deaths; EF term HR 2.50, 95% CI 1.49–4.21, P<0.001)** | | | | | | |
| Synthetic e′ (per 1 cm/s) | 0.85 (0.76–0.94) | 0.002 | 9.73 | 44% | 0.668 | +0.054 |
| Synthetic s′ (per 1 cm/s) | 0.81 (0.68–0.96) | 0.016 | 5.24 | 30% | 0.656 | +0.041 |
| Synthetic MAPSE (per 1 cm) | 0.58 (0.37–0.92) | 0.021 | 5.18 | 30% | 0.651 | +0.037 |
| Synthetic a′ (per 1 cm/s) | 1.06 (0.90–1.24) | 0.500 | 0.45 | 4% | 0.627 | +0.013 |

Each row is a Cox model containing the ejection-fraction term named in the section heading together with the single synthetic index named in the row. Hazard ratios are per 1 cm/s, or per 1 cm for MAPSE, so a value below 1 denotes higher mortality at lower values of the index. The likelihood-ratio chi-square is the contribution of the synthetic index on 1 degree of freedom, given also as a percentage of the full model chi-square. Delta C is the change in Harrell C-index relative to the ejection-fraction term alone. The three AI-ECG models were dichotomized at a predicted probability of 0.5. Synthetic a′ is pre-specified negative control and adds nothing in any of the four comparisons. The very large percentage shares in Section C are an artefact of a non-prognostic ejection-fraction term rather than a stronger synthetic signal: the HeartWise term itself carries a hazard ratio of 1.47 (95% CI 0.80 to 2.71, P=0.218), so almost all of that model chi-square must come from the added index. Section C is therefore excluded from the 28 to 46% range quoted in the main text. Section A is fit in 1,016 patients rather than the 1,017 of the full post-discharge cohort because one patient has no AiTiA prediction; Section D is fit in 997, the number with a measured echocardiographic ejection fraction. This table is the numerical basis of Figure 4 Panel A. Abbreviations: CI, confidence interval; df, degrees of freedom; EF, ejection fraction; HR, hazard ratio; LR, likelihood ratio; LVEF, left ventricular ejection fraction; MAPSE, mitral annular plane systolic excursion.

**Supplemental Table 13. Two-Year Kaplan-Meier Mortality by Predicted Ejection Fraction and Synthetic e′ Tertile (Post-Discharge AMI Cohort)**

| Group | e′ tertile | N | Deaths | 2-yr mortality (95% CI) | P for trend |
|---|---|---|---|---|---|
| AiTiA: predicted LVEF ≥ 50% | T1 (lowest) | 244 | 18 | 8.7% (4.6–12.6) | 0.004 |
| | T2 | 293 | 13 | 5.8% (2.6–8.8) | |
| | T3 | 320 | 7 | 2.5% (0.4–4.5) | |
| AiTiA: predicted LVEF < 50% | T1 (lowest) | 92 | 13 | 15.4% (6.1–23.7) | 0.838 |
| | T2 | 45 | 6 | 16.3% (1.7–28.7) | |
| | T3 | 22 | 3 | 9.5% (0.0–21.2) | |
| AI-ECG HFrEF risk: predicted low risk | T1 (lowest) | 248 | 22 | 9.6% (5.3–13.7) | < 0.001 |
| | T2 | 284 | 12 | 5.5% (2.3–8.6) | |
| | T3 | 323 | 8 | 2.5% (0.4–4.5) | |
| AI-ECG HFrEF risk: predicted high risk | T1 (lowest) | 88 | 9 | 13.4% (4.3–21.6) | 0.678 |
| | T2 | 55 | 8 | 18.5% (4.4–30.6) | |
| | T3 | 19 | 2 | 11.1% (0.0–24.5) | |
| HeartWise: predicted LVEF ≥ 50% | T1 (lowest) | 264 | 25 | 10.6% (6.2–14.8) | < 0.001 |
| | T2 | 282 | 15 | 6.5% (3.1–9.9) | |
| | T3 | 309 | 8 | 2.9% (0.7–5.1) | |
| HeartWise: predicted LVEF < 50% | T1 (lowest) | 72 | 6 | 10.1% (1.9–17.5) | 0.686 |
| | T2 | 57 | 5 | 11.4% (1.2–20.5) | |
| | T3 | 33 | 2 | 3.2% (0.0–9.2) | |

Kaplan-Meier estimates at 730 days with Greenwood 95% confidence intervals. Tertile cut points are the full clinical AMI cohort boundaries, 5.89 and 8.33 cm/s, applied unchanged to the post-discharge cohort. P for trend is from a Cox model on the ordinal tertile score within each predicted-ejection-fraction group. These are the numbers plotted in Figure 4, Panel B, which shows the same three AI-ECG models; measured echocardiographic LVEF is not shown here and appears in Supplemental Tables 11, 12 and 14.

### Supplemental Table 14. Synthetic e′ Entered as Tertile Indicators Within Each Representation of Ejection Fraction (Post-Discharge AMI Cohort)

| Ejection-fraction prediction | T2 vs T1 | T3 vs T1 | LR $\chi^2$ (2 df) | % of model $\chi^2$ | P |
|---|---|---|---|---|---|
| AiTiA — predicted LVEF < 50% | 0.70 (0.39–1.25) | 0.40 (0.19–0.82) | 6.9 | 24% | 0.032 |
| AI-ECG HFrEF risk — predicted LVEF ≤ 40% | 0.67 (0.38–1.17) | 0.36 (0.17–0.76) | 8.3 | 40% | 0.016 |
| HeartWise — predicted LVEF < 50% | 0.61 (0.35–1.06) | 0.30 (0.15–0.62) | 12.4 | 90% | 0.002 |
| Measured echo LVEF < 50% | 0.67 (0.38–1.19) | 0.38 (0.18–0.79) | 7.5 | 38% | 0.023 |

Each row is a Cox model containing the ejection-fraction term plus synthetic e′ as two tertile indicators, with the lowest tertile as reference. Cut points are the full clinical AMI cohort boundaries applied unchanged. The likelihood-ratio chi-square is the contribution of the two tertile indicators on 2 degrees of freedom, expressed also as a percentage of the full model chi-square. The 2-df tertile test is significant for every representation, with the highest tertile carrying roughly one third the mortality hazard of the lowest. The per 1 cm/s estimates in Supplemental Table 12 remain the more efficient test.

**Supplemental Table 15. Incremental Discrimination Beyond the AI-ECG HFrEF Risk Model for MI-Specific Mortality (Population Cohort, N=219,564)**

| Model | C-index (95% CI) | Delta C (95% CI) | P |
|---|---|---|---|
| AI-ECG HFrEF risk alone (binary at 0.5) | 0.525 (0.507–0.544) | - | - |
| + Synthetic e′ | 0.614 (0.595–0.633) | +0.089 (+0.066 to +0.112) | <0.001 |
| + Synthetic s′ | 0.586 (0.563–0.609) | +0.060 (+0.039 to +0.082) | <0.001 |
| + Synthetic MAPSE | 0.657 (0.637–0.677) | +0.132 (+0.108 to +0.155) | <0.001 |
| + Synthetic a′ | 0.526 (0.503–0.549) | +0.001 (–0.019 to +0.020) | 0.940 |
| + Synthetic s′ and MAPSE | 0.660 (0.640–0.680) | +0.134 (+0.111 to +0.158) | <0.001 |

Discrimination is the Wolbers competing-risk concordance index for MI-specific mortality, in which competing (non-MI) deaths are treated as non-events rather than as censored observations. The comparator is the AI-ECG HFrEF risk model of Pan et al., which predicts incident LVEF of 40% or less over a 1-to-5-year horizon, dichotomized at a predicted probability of 0.5; it was developed to forecast incident heart failure rather than death, which is why it discriminates only marginally for this outcome. Confidence intervals for Delta C come from paired concordance influence functions, so that each comparison is made within the same participants. Synthetic a′ is the pre-specified negative control and adds nothing (Delta C +0.001, P=0.940), indicating that the gains for e′, s′ and MAPSE are not an artefact of adding any continuous variable to a weakly discriminating model. CODE-15 contributed to neither AiTiA nor the model of Pan et al., so this comparison is external. Abbreviations: CI, confidence interval; LVEF, left ventricular ejection fraction; MAPSE, mitral annular plane systolic excursion; MI, myocardial infarction.

### Supplemental Figures

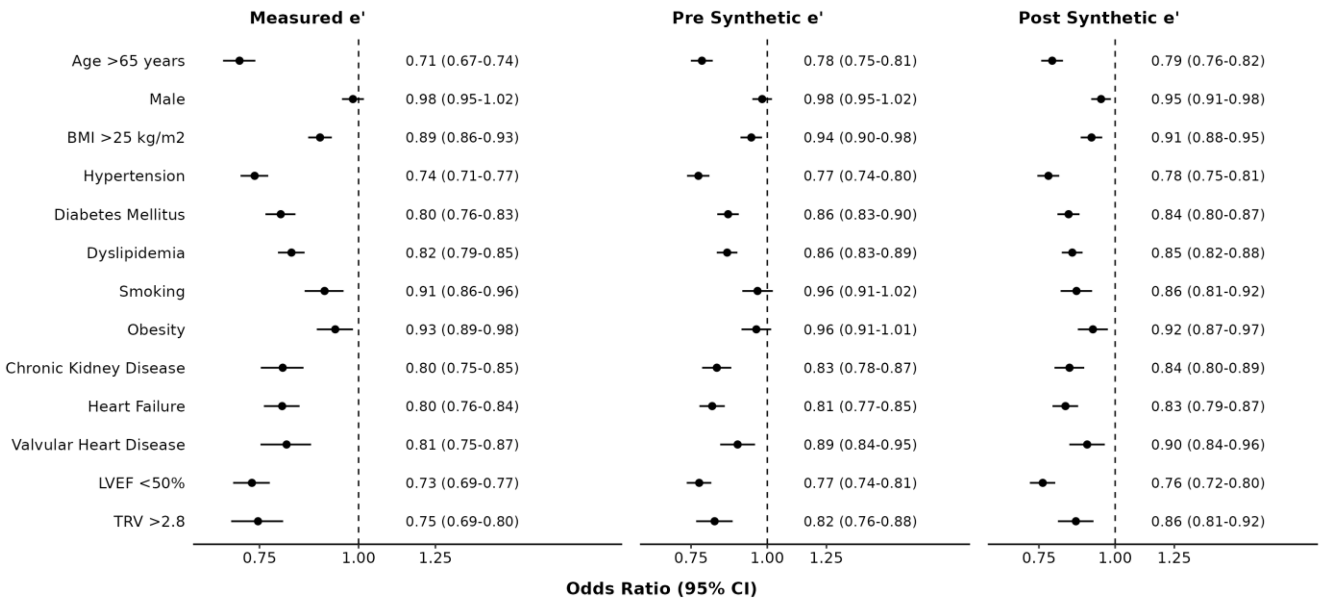


**Supplemental Figure 1. Odds ratios for cardiovascular risk factors per unit increase in synthetic and measured early diastolic mitral annular velocity.** Univariate logistic regression models fit with each cardiovascular risk factor as the binary outcome and synthetic e′ or measured e′ as the continuous predictor. Both synthetic e′ derived from pre-procedure ECGs and from post-procedure ECGs are shown. Results are expressed as odds ratios (ORs) with 95% confidence intervals. Models fit in the mixed clinical cohort of hospitalized AMI patients and outpatient non-AMI controls (n=1,749). The analytic sample reflects the paired-ECG restriction, namely the 803 AMI patients with both a pre- and a post-procedure ECG together with the 946 non-AMI controls and is not a consequence of missing data. Rows differ in the number analyzed because some characteristics were not recorded in every patient; tricuspid regurgitation velocity was unrecorded in 640 of 1,065 AMI patients and 329 of 946 controls, so that row rests on roughly half the sample. Across the thirteen rows the number analyzed ranges from 900 to 1,749: the measured e′ column is fit in 1,614 participants for most rows and the synthetic columns in 1,749, while the tricuspid regurgitation velocity row is the smallest, at 900 for measured e′ and 920 for the synthetic columns.
